# A Global Atlas of Immune Alleles Reveals Health-Relevant Variation Across Groups

**DOI:** 10.64898/2026.11.02.26367118

**Authors:** William T. Harvey, Anne-Katrin Emde, Fredrick E. Kakembo, Iman Hamid, Wayne E. Clarke, Isabella Apriyana, Hiba Babiker, Rindra Rakotoarivony, Melissa Hendershott, Prathap Kasina, Allan Kalungi, Oyesola Ojewunmi, Tristan Pascart, Stefania Rosso, Ricardo A. Verdugo, Catterina Ferreccio, Alain Dessein, Pradiptajati Kusuma, Lidwina Priliani, Safarina G. Malik, June Fabian, Michèle Ramsay, Kaja A. Wasik, P. Keolu Fox, Tony R. Merriman, Segun Fatumo, Herawati Sudoyo, Laura Yerges-Armstrong, Stephane E. Castel

## Abstract

Immune allele frequencies inform disease association mapping, pharmacogenomic guidance, and vaccine design, yet such data are missing for most of the world’s groups. Here we present the Global Atlas of Immune Alleles (GAIA), applying benchmarked genotyping tools across the Human Leukocyte Antigen (HLA), Immunoglobulin Heavy Chain Constant (IGHC), and Fc Gamma Receptor (FCGR) loci to whole-genome sequencing data from 20,248 individuals, oversampling sub-Saharan African, Island Southeast Asian, and Pacific groups. Despite representing fewer than 0.5% of observations in the Allele Frequency Net Database, GAIA expands the database’s catalogue by 38%, demonstrating that representation matters more than sample size. We identify HLA alleles at group-specific frequencies that challenge pharmacogenomic screening recommendations and document disease-risk alleles common in groups lacking clinical guidance. Finally, we introduce majority allelic diversity (MAD-X), which counts the alleles required for X% of individuals in a group to carry at least one, and show that reaching 90% HLA-B coverage for vaccine design requires up to 2.7 times more alleles in underrepresented groups than in those of Northwest European ancestry.

## Introduction

Immune response receptor genes are among the most polymorphic in the human genome. These include the classical human leukocyte antigen (HLA) genes, which have tens of thousands of allelic variants ^1, 2, 3^, and the immunoglobulin heavy chain constant region locus (IGHC), which contains dozens of highly similar, short gene segments, each with multiple alleles ^4, 5, 6^. This diversity is functionally essential, enabling recognition of diverse pathogens, determining immune cell isotypes, and influencing outcomes in infection, autoimmunity, and vaccine response ^7, 8, 9^. Specific HLA alleles affect drug reactions and disease susceptibility. For example, HLA-B*58:01 predisposes carriers to severe allopurinol reactions ^10^ and multiple HLA-B*27 subtypes are strongly associated with ankylosing spondylitis ^11, 12^. Fcγ receptors (FCGR) modulate antibody binding and effector functions, impacting immunity and therapeutic responses ^13^. Characterizing group-level immune gene variation is therefore critical for global health applications, particularly T cell vaccine design, where candidate peptides must bind HLA molecules expressed in the target group to elicit protective responses ^14^.

Despite their importance, these loci have been challenging to study at scale. Standard short-read variant calling does not adequately capture their diversity due to high sequence homology, tandem repeats, and structural variation ^15, 16^. Global resources such as the 1000 Genomes Project (1kGP) include diverse groups but remain limited in scope and rely on standard short-read pipelines, resulting in as much as ∼20% of common and ∼70% of rare HLA alleles being absent from the 1kGP dataset due to limited group sampling, underscoring the incompleteness of current immunogenetic catalogues ^17^.

Specialized bioinformatics tools have been developed to infer genotypes at these complex loci from short-read whole-genome sequencing (WGS) data ^18, 19, 20, 21, 22, 23^. In parallel, large-scale long-read sequencing efforts by the Human Genome Structural Variation Consortium (HGSVC) and Human Pangenome Reference Consortium (HPRC) have produced sequence-resolved assemblies that improve our understanding of immune receptor variation and provide diverse reference material for benchmarking. Together, these advances create an opportunity to systematically map immune receptor variation at scale using existing WGS datasets.

Here we present the Global Atlas of Immune Alleles (GAIA), the largest map of immune receptor gene allele frequencies to date. We analyzed WGS data from over 20,000 individuals across Africa, Asia, the Americas, Oceania, and Europe, with particular emphasis on sub-Saharan Africa (n = 6,163), Island Southeast Asia (n = 2,536), and the Pacific Islands (n = 4,095). After benchmarking specialized genotyping tools against haplotype-resolved reference assemblies, we calculated allele frequencies stratified by geographic groups and identify alleles whose frequencies diverge substantially from the Allele Frequency Net Database (AFND), or for which we provide the first frequency estimate ^1^. To quantify how immune diversity is distributed within groups, we introduce majority allelic diversity (MAD-X), defined as the number of alleles selected by a greedy algorithm until at least X% of individuals in a group carry at least one. Finally, we highlight clinically relevant variation in groups where it has not previously been well-characterized, demonstrating the concrete health consequences of the diversity gaps revealed by GAIA.

## Results

### GAIA Study Description and Tool Benchmarking

To profile immune receptor allele frequency at a global scale, we assembled a diverse cohort of individuals with short-read WGS from multiple publicly available studies and newly generated data from 8,463 individuals (see Methods). We recognize that human genetic variation is continuous and does not fall neatly into discrete groups. However, in order to describe the global distribution of allele frequencies for the purposes of this study, we grouped individuals by continental (e.g. Africa, Oceania, Europe, etc.) and subcontinental (e.g. Northern, Western, Eastern, etc.) regions and calculated allele frequencies within these groupings (see Supplemental Table S1 for group definitions). After quality control (QC) filters (see Methods), the final GAIA dataset included 20,248 individuals from 18 subcontinental regions (Figure 1A, B, Table S1). Sample sequencing coverage ranged from 4× to 66× with a bimodal distribution of 11,330 samples below 20× (Mean: 6.2×, Median: 5.1×) and 8,918 above 20× (Mean: 37.7×, Median: 36.3×) (Supplemental Figure S1).

**Figure 1.**
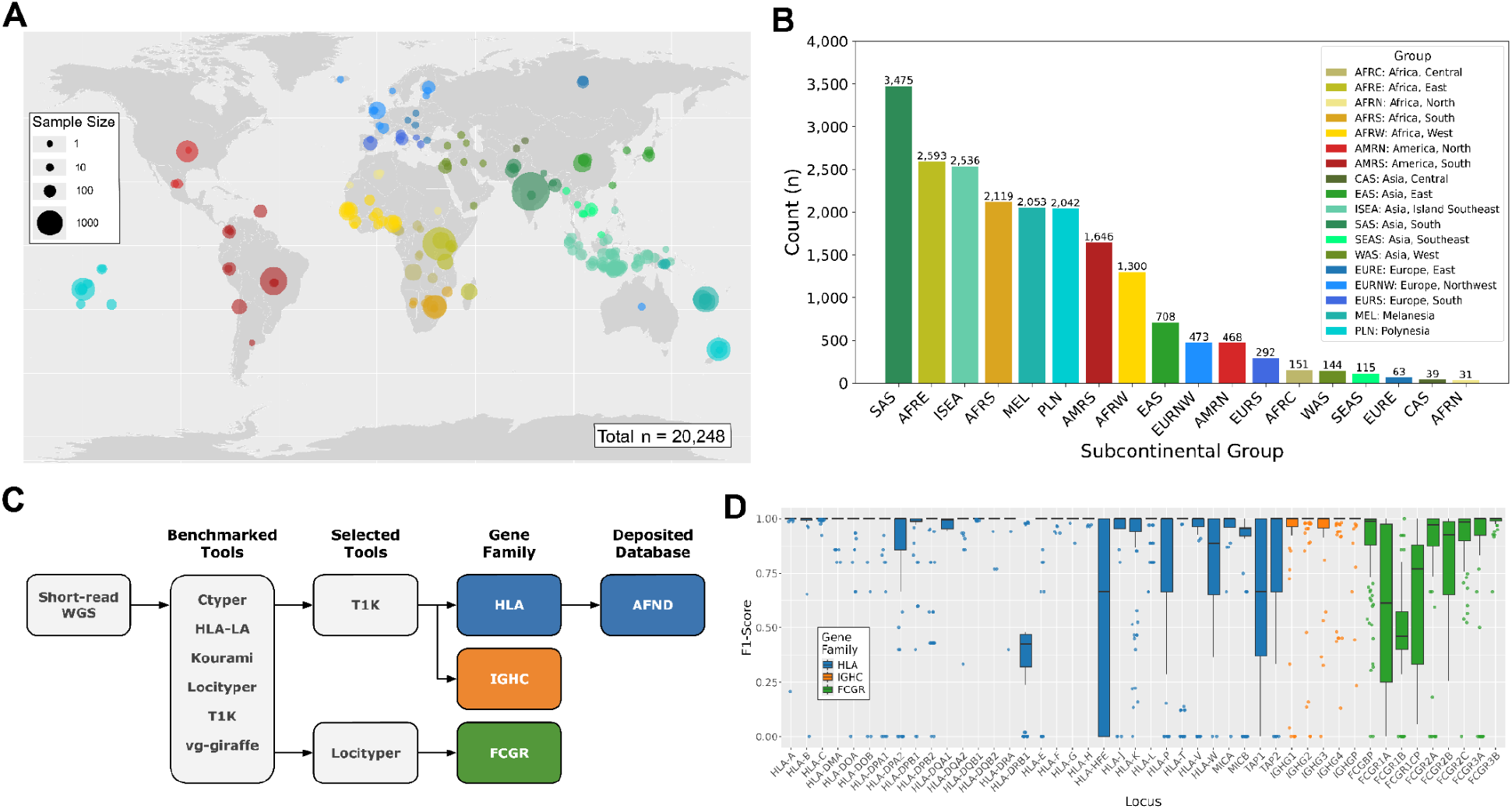
The Global Atlas of Immune Alleles (GAIA) study. A) Groups sampled in GAIA plotted across a world map with marker size correlating to sample size and color corresponding to the subcontinental group. Multiple continental groups can be incorporated in the same subcontinental group as reported in the manuscript (Supplemental Table S11). B) Overall sample counts per subcontinental group. C) Analysis workflow of input data and genotyping tools per immune gene family. Following extensive benchmarking of multiple tools, T1K (HLA, IGHC) and Locityper (FCGR) were selected for downstream use. D) F1-scores for 55 samples from the 1000 Genomes Project downsampled to 10× depth, comparing predicted genotypes against haplotype-resolved long-read assemblies. Scores were computed from GRCh38 alignments using hap-eval (see Methods). Box plots colored by gene family: HLA (blue, T1K), IGHC (orange, T1K), FCGR (green, Locityper).

Accurate genotyping of HLA, IGHC, and FCGR required specialized bioinformatics tools, which we selected through systematic benchmarking against haplotype-resolved assemblies and high-confidence reference data (Figure 1C–D). This benchmarking showed that T1K^19^ achieved the best performance for HLA and IGHC, while Locityper^18^ performed best for FCGR. Genotyping accuracy was high for HLA and IGHC across all coverage depths tested, with F1-scores exceeding 0.89 at 5× sequencing depth; FCGR was somewhat harder to genotype accurately, with an F1-score of 0.79 at 5× depth (Supplemental Table S2). We report an overall score per locus, though accuracy varies substantially between genes; that breakdown, along with an estimation of the effect of sequencing depth on the number of alleles with frequencies reported for the first time (Supplemental Figure S2), is detailed in the Methods.

### HLA Frequency Estimates Diverge from Current References

We compared HLA allele frequencies in GAIA with those reported in the AFND to assess how allele frequencies in historically underrepresented groups differ from those currently catalogued. For each identified allele, we computed a GAIA-wide frequency, compared it with the corresponding AFND frequency, and identified the most differential alleles. Twenty-four HLA alleles, which were previously reported in the AFND, had study-wide frequencies that differed from AFND values by more than 5% (Figure 2A, Table S3). Two of the most abundant alleles in the AFND, A*01:01 (13.1%) and A*02:01 (23.9%), occurred at substantially lower frequencies in GAIA (5.1% [95% CI: 4.9-5.3%] and 9.2% [95% CI: 8.9-9.4%] respectively). Alleles in the A*01 group have been linked to human immunodeficiency virus (HIV) susceptibility^24^ although A*01:01 specifically has been associated with better protection from malaria after RTS,S vaccination^25,26^. C*07:01, which is associated with HIV susceptibility^24^, occurred at ∼7% (95% CI: 6.5-7.4%) frequency in the continental African subset of GAIA, yet is reported at nearly twice that frequency (13.4%) in the AFND. A*24:02 has been associated with greater susceptibility to type 1 diabetes (T1D)^27,28^ and was observed in GAIA at a frequency of 15.3% (95% CI: 14.9-15.6%), 6% higher than that reported in the AFND (9.3%).

**Figure 2.**
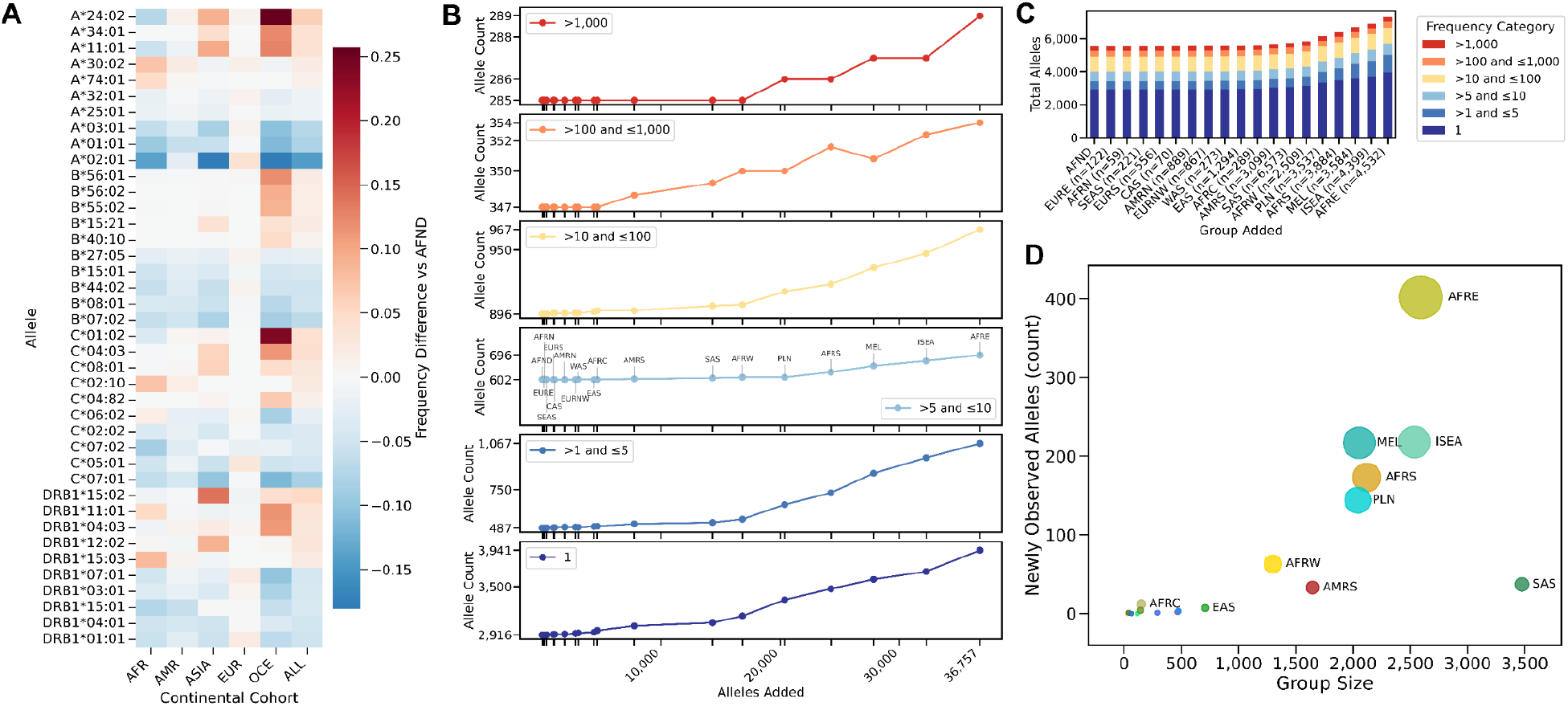
Comparison of GAIA HLA Allele Frequencies to AFND. A) Heatmap demonstrating the 5 most positive and 5 most negative differential allele frequencies per locus between the AFND and allele frequencies reported in GAIA (minimum 75 occurrences in GAIA). Alleles were selected based on global differential and split out by continental group. B) Newly observed allele and allele frequency change in HLA-A, -B, -C, and DRB1 for various observation frequency categories in the AFND after adding each group individually and plotted by group size to the database. The middle subpanel for frequencies >5 and ≤10 is annotated with the populations in the order in which they were added. This order is the same across all subpanels in panel C. C) Cumulative allele counts for HLA-A, B, C, and DRB1 for each frequency category after adding groups in the same order as panel B. The first bar represents the current status of the AFND and the last bar represents the status of the AFND after addition of GAIA frequencies. D) Number of newly observed alleles unique to a given subcontinental group by group size. Newly observed alleles have sequences in the IPD-IMGT/HLA but no allele frequencies in the AFND and are unique to specific subcontinental groups are represented by circles on the scatter plot and measured on the y-axis. Circles are sized by the number of alleles which are unique to only that subcontinental group as an additional demonstration. The x-axis tracks the number of individuals in the subcontinental group.

Despite GAIA constituting fewer than 0.5% of the samples in the AFND (n = ∼7 million with frequency information for A, B, C, and DRB1), it reports allele frequencies for 2,413 alleles for the first time, which cumulatively increase the number of alleles with frequency information in the AFND by 38% (Table S4). Of the alleles with newly reported frequencies, 1,780 are in the most well-characterized subcomplexes (HLA-A, -B, -C, and -DRB1 [Table S5]) and 1,007 alleles were observed more than once in GAIA (Table S4). These 1,780 contained an allele from each subcontinental group with the exceptions of Eastern Europe and Northern Africa which were populated solely by samples from reference cohorts. Projections indicate that the discovery of previously unreported allele frequencies is still accelerating with each new individual added to the study (Figure 2B-C). Of the 2,413 previously unreported alleles, 20 had an observed allele frequency greater than 1% (Table S6) in a given subcontinental group, one of which (DQB1*04:59) was detected at a study-wide frequency greater than 1% (3.5% [95% CI: 3.3-3.6%]). With the exception of South Asia and Southeast Asia, every subcontinental group surveyed contained an example of an allele that was previously unreported and occurred at greater than 1% frequency in that group, but this pattern was most frequently observed in Polynesia (6 alleles), Central Asia (6 alleles), Eastern Africa (5 alleles), and Central Africa (4 alleles). 1,317 of the 1,780 alleles in the HLA-A, -B, -C, and -DRB1 subcomplexes that were not observed in the AFND were unique to specific subcontinental groups (Figure 2D), with the largest numbers of new observations coming from Eastern Africa (402), Island Southeast Asia (218), and Melanesia (217). In addition to these alleles, for which frequency information is reported for the first time, GAIA provided frequencies for 583 alleles across HLA-A, -B, -C, and -DRB1 that previously had been reported in the entire AFND fewer than 5 times. These results highlight how even modest increases in global representation in allele discovery can uncover substantial novel frequency information.

### Global Frequencies of IGHC and FCGR Alleles

Although the largest portion of genetic immunology research has centered on HLA, the immune response is dictated by a host of genes and mechanisms. Genes belonging to the IGHC and FCGR families have been implicated in disease-related outcomes^29,30,31,32^ but their diversity is less well documented and cataloged. While some efforts to catalog and describe IGHC alleles have been successful^33,34^, no database exists for cataloging FCGR. The absence of these efforts can be seen in the benchmarking results where the genotyping accuracy for genes within these families is generally lower than what is observed in the major HLA subcomplexes and in the smaller number of alleles used in our genotyping databases (average 55 per IGHC gene vs 1,180 per HLA gene). Despite this limitation, we note large differences in the most abundant allele per subcontinental group for the IGHC genes.

IGHG1*02 is the most abundant allele in the European (56.5% [95% CI: 53.8-59.3%]) and American (38.9% [95% CI: 37.2-40.7%]) continental groups, and is present at 23.1% (95% CI: 22.3-24.0%) in Asian groups, where IGHG1*05 is more abundant (28.8%) (Supplemental Figure S3). It occurs at 7.4% (95% CI: 6.7-8.1%) in Oceanian and 1.2% (95% CI: 1.0-1.4%) in African groups. For FCGR, East Asia, Melanesia, Northwest Europe, Polynesia, South Asia, and Southeast Asia showed an increase in alleles originating from 1kGP samples of Western African ancestry over the FCGR1 gene, and in most cases it was the most common graph-based haplotype of origin (Supplemental Figure S4, Methods). This pattern was not observed across FCGR2 or FCGR3 genes. Some technical factors could complicate the analysis: FCGR1A and FCGR1B have the lowest benchmarking performance in the FCGR locus, and this allele set is drawn specifically from alleles in the HPRC graph, so the signal could stem from underrepresentation of Pacific Islander individuals in the graph. The significance of the results for IGHC and FCGR are not yet understood, but we report them to encourage future research.

### Majority Allelic Diversity Quantifies the Cost of Equitable Vaccine Design

Understanding global diversity has a direct impact on health initiatives, not least of which is vaccine design. Achieving broad protective coverage and evaluating safety in epitope-based vaccine design typically requires targeting alleles sufficient to cover at least 90% of individuals in a group^35^, with earlier work establishing 80% coverage from 3-5 molecules per ethnicity but requiring different specificities in each^36^; population coverage is conventionally computed from HLA genotype frequencies and epitope restriction data^37^. Existing diversity metrics summarize variation at a locus as a single value^38^, but do not directly indicate how many alleles must be included to reach a given level of population coverage. To make such comparisons tractable, we introduce majority allelic diversity (MAD-X), defined as the number of alleles selected by a greedy algorithm until at least X% of individuals in a group carry at least one. MAD provides a direct measure of how concentrated or dispersed allelic diversity is: at lower thresholds (MAD-50) it quantifies how few alleles suffice to reach half of a group, and at thresholds relevant to vaccine design (MAD-90) it quantifies the breadth of alleles required to achieve broad group coverage.

To illustrate this point, we focus on the HLA-B locus which shows the highest MAD-50 among the HLA genes with a mean of 4.8 between continental groups (Figure 3A), consistent with prior findings that HLA-B is the most diverse HLA subcomplex and with evidence that HLA-B dominates CD8+ T cell recognition and immune selection where locus-specific contributions have been directly measured^39,40,41^. While MAD-50 captures where common diversity is concentrated, MAD-90 therefore provides a direct quantification of the allelic breadth required for equitable vaccine design at each locus and in each group. The HLA-B locus had a mean MAD-90 of 16.2 between continental groups, followed by DRB1, C, and A with mean MAD-90 values of 10.8, 9.2, and 8.4 respectively (Figure 3B).

**Figure 3.**
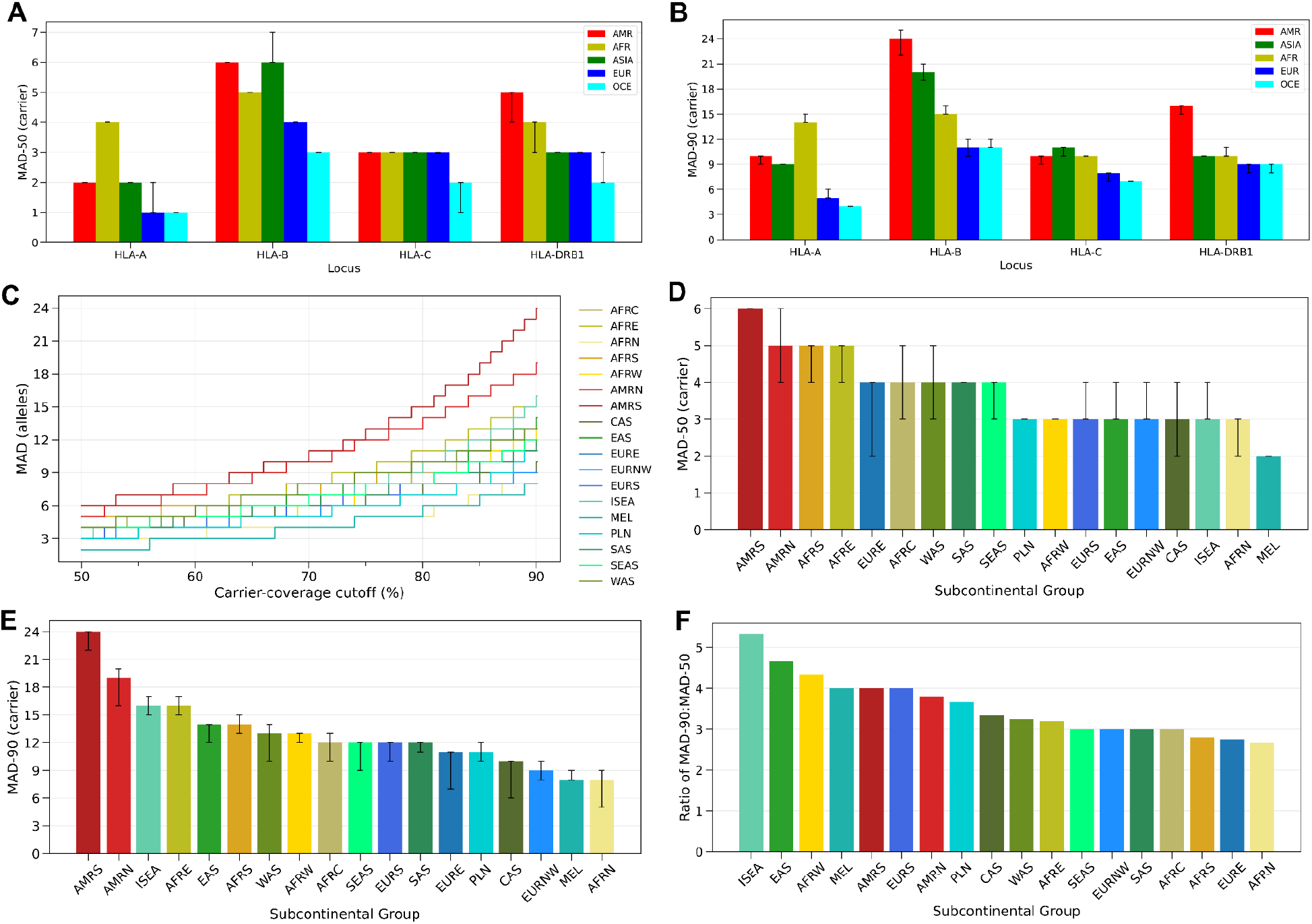
Majority allelic diversity (MAD) quantifies the allelic breadth required for group coverage. A-B) Number of alleles to ensure 50% (MAD-50, A) or 90% (MAD-90, B) of individuals in a group carry at least one allele from the set across HLA-A, -B, -C, and -DRB1 for each continental group. C) Plot of MAD-X for each subcontinental group at HLA-B. Lines are colored by subcontinental groups. X-axis indicates the MAD threshold and the y-axis is the number of alleles required to reach that threshold. D) MAD-50 for all subcontinental groups at HLA-B. E) MAD-90 for all subcontinental groups at HLA-B. F) Ratio of HLA-B MAD-90 to MAD-50 for each subcontinental group. This ratio represents the relative number of alleles required to ensure 90% of individuals in a group have at least one allele from the set compared to the set for 50% of individuals. NOTE: Error bars for D & E represent 95% confidence intervals based on 1,000 bootstraps of sampling with replacement from the alleles in each group up to the group size.

Achieving 90% HLA-B coverage at the subcontinental level required 9 alleles (95% CI: 8-10) in the Northwest European group compared with 16 alleles (95% CI: 15-16; 1.78-fold higher) in the Eastern Africa group and a maximum of 24 (95% CI: 22-24; 2.7-fold higher) in the South American group, with similar but smaller gaps observed at HLA-A, -C, and -DRB1 (Figure 3C-E, Supplemental Figure S5, Table S7). To achieve 90% coverage in every subcontinental group, the HLA-B locus required 61 alleles compared to 31, 27, and 35 alleles at HLA-A, -C, and -DRB1, respectively. The ratio of MAD-90 to MAD-50 provides a measure of how heavy the tail of common variation is at each locus. For HLA-B, across subcontinental groups, this ratio spanned from 5.3 in Island Southeast Asia to 2.7 in Northern Africa, revealing that the alleles required for broad coverage outnumber those that account for the bulk of the group (Figure 3F). These findings indicate that the operational cost of designing globally equitable T cell vaccines is concentrated at HLA-B and falls disproportionately on groups whose diversity is least represented in current reference databases.

### Health Implications of Group-Specific HLA Variation

We surveyed HLA alleles with known impacts on clinical and global health outcomes (Supplemental Table S8, S9) and identified those occurring at common frequency (>1%) in specific subcontinental groups. Organizing these findings by clinical actionability revealed three categories: (1) confirmatory observations that validate established group-allele patterns at unprecedented resolution, (2) alleles indicating that current pharmacogenomic guidance may be miscalibrated for specific groups, and (3) disease-associated alleles common in groups where they have not previously been well-characterized (Figure 4, Supplemental Table S10).

**Figure 4.**
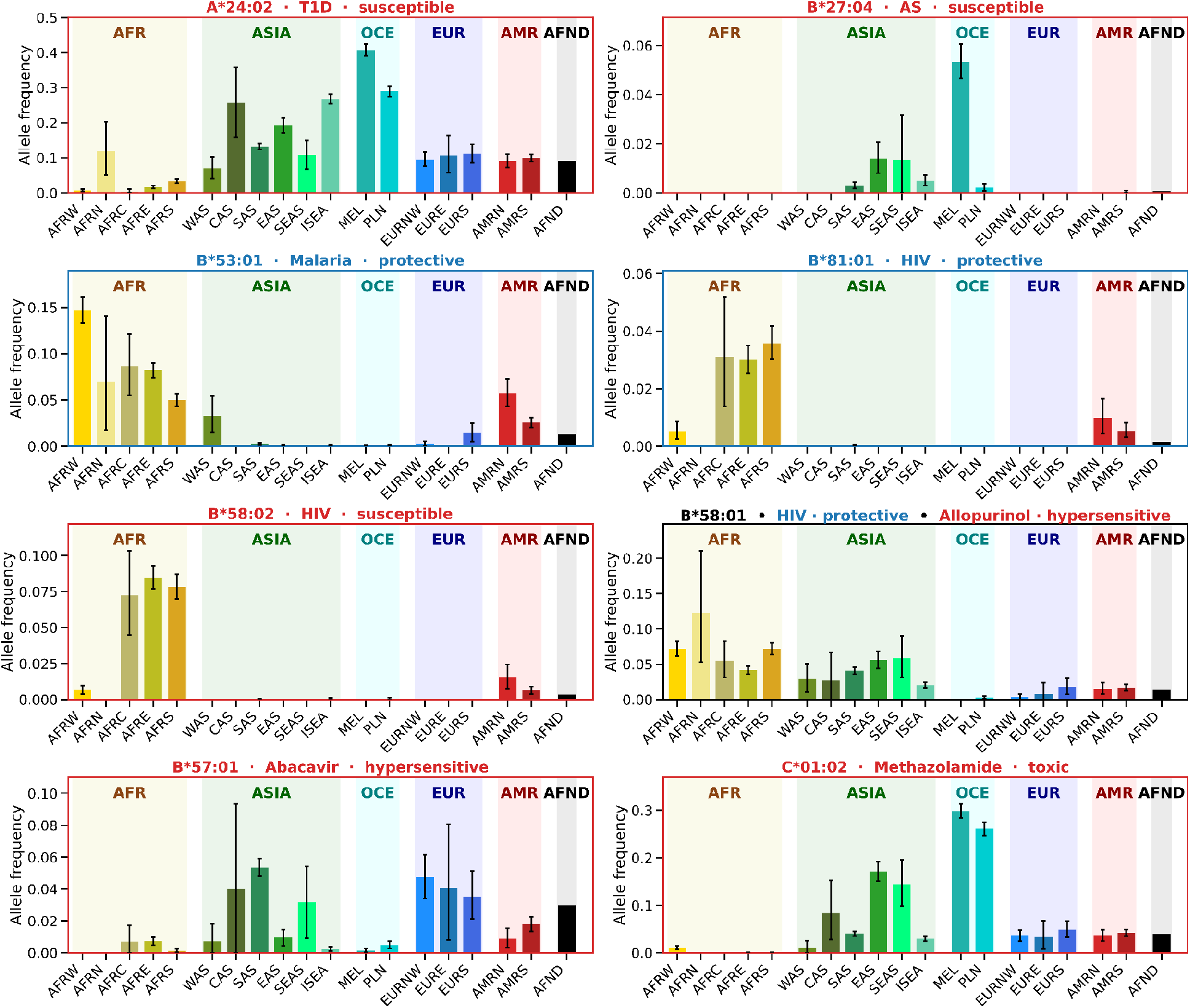
Allele frequencies of health-relevant HLA alleles for each subcontinental group. Allele frequency plots are color-coded by clinical effect (red = risk allele, blue = protective allele). These alleles encompass effects on diseases: malaria, type 1 diabetes (T1D), ankylosing spondylitis (AS), and human immunodeficiency virus (HIV) as well as adverse responses to pharmaceutical compounds: abacavir, allopurinol, and methazolamide. Variable frequencies represent variable risks in each subcontinental group. Error bars represent 95% confidence intervals based on 1,000 bootstraps of sampling with replacement from the alleles in each group up to the group size.

#### Validation of established group-allele patterns

GAIA recapitulates a number of well-established group-specific allele distributions at a larger scale than originally reported. The African continental group carries a group of HLA-B alleles known to modulate HIV-1 disease outcomes, particularly for the subtype C virus that predominates in sub-Saharan Africa. These include B*57:03, the class I allele most strongly associated with suppression of HIV without the use of antiretroviral therapy^42,43,44,45^; B*81:01 and B*58:01, which mediate protection through Gag-restricted CD8+ T cell responses^46,47,48,49,50^; and B*42:01, which is genetically related to B*81:01 but confers more modest protection^51^. The protective B*57:02 subtype was observed at greater than 1% frequency in the Eastern African group (1.4% [95% CI: 1.0-1.7%]). Conversely, B*58:02, which is associated with rapid disease progression due to its failure to mount effective Gag-specific immune pressure^52^, is also common only in the African continental group, illustrating that common alleles in this region include those that are both protective and risk-associated (6.4% [95% CI: 5.9-6.8%]). Another disease-specific allele, B*53:01 which is associated with protection from severe malaria^53^, is common across Africa, ranging from 5.0% in Southern Africa (95% CI: 4.3-5.7%) up to 14.7% in Western Africa (95% CI: 13.3-16.1%). The Asian continental group carries B*46:01 at 1.5% (95% CI: 1.3-1.7%), associated with nasopharyngeal carcinoma in Chinese groups^54^, and the American continental group carries B*39:06 at 1.0% (95% CI: 0.7-1.4%), linked to accelerated HIV progression^55,56^. These confirmatory observations validate GAIA’s recovery of clinically relevant variation and provide the largest-scale frequency estimates to date for these alleles in the groups where they are of high importance.

#### Group-specific frequencies refine pharmacogenomic guidance

Pharmacogenomic screening recommendations often use geography or diaspora-based characterization and may benefit from more comprehensive characterization of allele frequencies in GAIA. Two examples illustrate how this can both under- and over-estimate the group frequency of clinically-actionable alleles. The Clinical Pharmacogenetics Implementation Consortium (CPIC) recommends HLA-B*57:01 testing in all abacavir-naive patients^57^, but B*57:01 is not detected in Western and Northern African groups in GAIA, consistent with published evidence that universal screening thresholds may not be warranted in these groups^58^. In contrast, the 2020 American College of Rheumatology (ACR) guideline recommends HLA-B*58:01 testing in patients of Korean, Han Chinese, or Thai descent, with additional consideration for African American patients for allopurinol hypersensitivity syndrome^59^, but GAIA subcontinental group-level frequency data reveal meaningful gaps. B*58:01 reaches 12.3% (95% CI: 5.3-21.1%) in Northern African and 7.2% in Western and Southern African groups (95% CI: 6.2-8.2% and 6.3-8.0% respectively) which is substantially higher than the African American estimates that informed the guideline (3.8%) and suggests that framing the African ancestry recommendation around diaspora groups underestimates risk for patients from continental Africa. Conversely, B*58:01 is essentially absent in Polynesian groups (0.27% [95% CI: 0.11-0.46%]) despite geographic proximity to screened East Asian groups, consistent with recent evidence supporting allopurinol prescribing in Tahitian patients without HLA screening^60^.

Although structurally similar to HLA-B*15:02, which forms the basis for carbamazepine hypersensitivity screening in East and Southeast Asia, the B*15:08 and B*15:11 alleles are potentially common in neighboring groups that current screening guidance does not cover. B*15:08 was observed at greater than 1% frequency in the Central Asian group (1.3% [95% CI: 0.0-4.0%]) and B*15:11 at greater than 1% frequency in the Southeast Asian group (1.8% [95% CI: 0.5-3.6%]). Both share the structural features that enable carbamazepine binding and may confer similar SJS/TEN risk^61^. Emerging pharmacogenomic associations also warrant attention to group frequency before guidance recommendations are established or developed. HLA-B*59:01 and HLA-C*01:02 have been identified as risk alleles for methazolamide-induced SJS/TEN with 2A and 2B evidence levels in ClinPGx^62^. HLA-B*59:01 itself is rare across most groups, which may limit the practical case for screening. While HLA-C*01:02 has a <5% frequency in European groups, it reaches 17.1% (95% CI: 15.1-19.2%) in East Asian, 26.1% (95% CI: 24.7-27.5%) in Polynesian, and 29.8% (95% CI: 28.4-31.3%) in Melanesian groups. This would be of major clinical consequence if the strength of the association evidence clears a higher evidence threshold and could also inform which groups are best suited for future confirmatory studies.

#### Disease-associated alleles in groups where they have not been well-characterized

B*27:04, one of the most strongly disease-associated B*27 subtypes for ankylosing spondylitis (AS) and related spondyloarthropathies^63,64^, was observed at 5.3% (95% CI: 4.6-6.1%) frequency in the Melanesian subcontinental group. B*27:04 is most commonly associated with East and Southeast Asian groups^65,66^, and its presence at common frequency in Oceania (2.8% [95% CI: 2.4-3.2%]), specifically Melanesia, has not been well-documented in prior studies. B*27:06, however, is a B*27 subtype that is notably not associated with AS, making it clinically important for distinguishing pathogenic from benign B*27 results, and occurs at greater than 1% frequency in Island Southeast Asia (3.3% [95% CI: 2.7-3.8%]). The combined picture across these regions demonstrates that B*27 typing alone is insufficient for AS risk stratification in Asia–Pacific groups; subtype resolution is required, and the relevant subtypes differ by group. A*24:02, an allele independently associated with increased risk and earlier onset of clinical T1D^27,28^, is present at exceedingly common frequencies in the Melanesian and Polynesian subcontinental groups, at 40.8% (95% CI: 39.1-42.4%) and 29.0% (95% CI: 27.5-30.5%) respectively, which substantially exceeds the allele frequencies observed in European groups (Northwest Europe: 9.6% [95% CI: 7.6-11.5%]; Southern Europe: 11.2% [95% CI: 8.6-13.8%]) and the AFND and surpasses even the elevated frequencies (17%) documented across mainland East Asia^67^. Supporting these findings, A*24:02 has previously been reported at high frequency in Oceania, including 37% among Māori^68^ and a mean allele frequency of 46.1% across Oceania and Southeast Asia, where it is the dominant ligand for inhibitory KIR3DL1 and carries evidence of positive selection^69^. T1D risk may be less recognized in Pacific Island communities, where background rates of type 2 diabetes are high^70,71^ and latent autoimmune diabetes occurs in 5.3% of Māori and Pacific adults diagnosed with type 2 diabetes^72^.

## Discussion

GAIA addresses a critical gap in immunogenomics by providing the most complete survey to date of immune gene loci and allele frequencies across genetically diverse and historically underrepresented subcontinental groups, and introduces majority allelic diversity (MAD) as a framework for quantifying the allelic breadth required for equitable group coverage. Together, the atlas and the metric reveal that current reference databases underestimate the diversity that vaccine design and pharmacogenomic guidance must accommodate, and that this underestimation disproportionately affects groups already underserved by precision medicine.

Our MAD-90 analysis quantifies this challenge directly: achieving 90% group coverage for vaccine development at HLA-B requires 61 alleles in the GAIA cohort overall, and up to 24 alleles in the South American subcontinental group, substantially more than would be inferred from current reference databases. Notably, the North and South American groups included many admixed individuals, which may contribute to the high MAD-90 values observed. Admixture averages allele frequencies across source populations weighted by their relative contributions, shifting alleles toward intermediate frequencies and away from fixation, and thus requiring more alleles to reach a given coverage threshold. The fact that MAD-90 exceeds MAD-50 at HLA-B by a factor of ∼2 globally and up to 5.3 on a group-by-group basis indicates that the alleles required for broad protective coverage extend well beyond those that dominate the group, and that this tail is most pronounced in the groups least represented in existing resources. HLA variation has been shown to drive measurable differences in vaccine antibody response in African groups^73^, underscoring that reference databases skewed toward well-studied groups risk producing vaccines with uneven global coverage. MAD-90 makes that risk quantifiable, and identifies HLA-B as the locus where the operational cost of equitable design is concentrated.

The atlas itself contributes a 38% increase in the number of alleles with frequency information in the AFND, while comprising fewer than 0.5% of the total observations in the database. This rate of discovery indicates that representation, not sample size alone, is the limiting factor for completing global immunogenetic catalogues, and that even modest expansions of the GAIA dataset are likely to reveal further novel allele frequency information for the remaining 30,651 (80.5%) IPD-IMGT/HLA^74^ classified alleles for which no group-level frequency data currently exist.

GAIA is a first step toward closing the gap in our understanding of how these alleles shape health outcomes and interact with one another and with environmental factors. GAIA offers a more equitable approach to allele frequency estimates but remains subject to sampling bias arising from study design (e.g., cohorts recruited under specific clinical conditions). While our MAD metrics quantify the scope of common immune diversity and reveal distinct patterns across less well-studied subcomplexes, linking specific alleles to functional outcomes in underrepresented groups will require further research that incorporates direct clinical or immunological measurements. MAD also counts alleles rather than epitopes; because individual peptides frequently bind multiple alleles within an allelic family or supertype, MAD-90 bounds the number of distinct binding specificities a vaccine must accommodate rather than the number of peptides it must contain^75^.

The clinical and genetic findings highlighted here focus on HLA alleles in part because the IGHC and FCGR loci lack a centralized resource analogous to AFND and IPD-IMGT/HLA, but also because the health implications of IGHC variation in particular remain less well-characterized than those of HLA despite similar allelic diversity^76^. Establishing a unified database for IGHC and FCGR would enable systematic comparison of allele frequencies across groups and, alongside continued functional study, provide a foundation for understanding these genes’ roles in disease susceptibility and treatment response.

While the derivation of these results from short-read WGS provides advantages over other methods, there is an inherent limitation to this technology specifically in high homology and copy number variable regions. New technologies and resources will continue to enable a more complete assessment of immune gene diversity. Long-read sequencing and high-fidelity assembly techniques can resolve complex loci and have revealed many novel alleles and structural haplotypes in IGH, IGK, and IGL loci^77,78^. For instance, a study of IGH^77^ identified 130 previously undocumented alleles not present in reference databases. Long-read assemblies from HGSVC have also demonstrated near-perfect HLA-typing while resolving previously incomplete HLA allele annotations and uncovering novel structural variation affecting genes such as HLA-DPA2^78^. These approaches will enable more complete characterization of complex genetic variation at these loci, including copy number and structural variation, and, coupled with richer health data, will improve our ability to link immune gene variation to health outcomes.

Understanding the full breadth of human immune diversity is not only an academic exercise in equity but a measurable prerequisite for global precision public health. MAD-90 provides one way to make that prerequisite concrete: it tells us how many alleles a vaccine, a screening guideline, or an association study must accommodate to serve a given group, and it shows that current reference databases systematically underestimate that number for most of the world. Closing that gap is a tractable problem, and GAIA demonstrates that even modest investments in representation yield substantial returns.

## Acknowledgements

First and foremost we would like to acknowledge the contributions of all participants who contributed data across the various cohorts. We also acknowledge the work of 1000 Genomes Project Consortium (1kGP), the Human Genome Diversity Project (HGDP), Indonesian Genome Diversity Project (IGDP), Papuan Genome Diversity Project (PGDP), Simons Genome Diversity Project (SGDP), Uganda Genome Resource (UGR), Human Heredity & Health in Africa (H3Africa), Malaria Genomic Epidemiology Network (MalariaGEN), Health, Well-being and Aging in São Paulo State, Brazil (SABE) Study, Sandra Beleza (University of Leicester, Angola and Mozambique study), Longitudinal Aging Study in India Diagnostic Assessment of Dementia (LASI-DAD), to generate data that were analyzed in this study. Data processing and analysis for this study was funded by the Gates Foundation grant INV-078742.

## Declaration of interests

WTH, AKE, IH, WEC, HB, MH, PrKa, KAW, PKF, TRM, LYA, and SEC were or are employees, advisors, and/or equity owners at Variant Bio. The remaining authors declare no competing interests.

## Author Contributions

AKE, LYA, and SEC designed the study. IA, RR, MH, PrKa, PrKu, TP, RV, CF, AD, SGM, JF, MR, KAW, PKF, TRM, SF, SR, AK, OO, LP, and HS were involved in the generation of data used in the study. AKE, SEC, and HB contributed to accessing public datasets. WTH, AKE, FEK, IH, and WEC performed data analysis. WTH, AKE, FEK, IH, LYA, and SEC drafted the manuscript, and all authors contributed to writing and reviewing the manuscript.

## Data Availability

HLA allele frequencies will be deposited to the AFND after peer review. All allele frequencies and supplemental tables are available on GitHub (https://github.com/variant-bio/gaia/), and large supplemental materials are available on Zenodo: https://doi.org/10.5281/zenodo.21515075. An interactive browser of allele frequencies and MAD-X is available at https://gaia.variantbio.com/.

## Code Availability

The custom scripts used for integrating genotyping results and performing analyses are available on GitHub (https://github.com/variant-bio/gaia/).

## Methods

### Cohort Description and Sample Preparation

This study aggregated whole-genome sequencing datasets from publicly available sets including the 1kGP^1^, Angola-Mozambique (University of Leicester)^2^ [EGAD00001011992]; Gambian Genome Variation Project^3^ [European Nucleotide Archive (PRJEB3013 (Fula), PRJEB3252 (Jola), PRJEB1682 (Mandinka) and PRJEB1323 (Wollof))], Human Genome Diversity Project (HGDP)^4^, Indonesian Genome Diversity Project (IGDP)^5^ [EGAD00001004156; EGAD00001005059], the Longitudinal Aging Study in India Diagnostic Assessment of Dementia (LASI-DAD)^6^, Papuan Genome Diversity Project (PGDP)^7^ [EGAD00001007783], Health, Well-being and Aging in São Paulo State, Brazil (SABE)^8^ [EGAD00001008640], the Simons Genome Diversity Project (SGDP)^9^, and the Ugandan Genome Resource^10^ [EGAD00001001639].

This study makes use of data generated by H3Africa^11,12^. A full list of the investigators who contributed to the generation of the data is available from https://h3africa.org. The funding for this project comes through the Human Heredity and Health in Africa (H3Africa) Initiative, which is funded by the National Institutes of Health and the Wellcome Trust through SFA Foundation. The following H3A datasets were included in the analysis: H3Africa H3AChipDesign TrypanoGEN2 (EGAD00001004220), H3Africa H3AChipDesign ELSI (EGAD00001004316), H3Africa H3AChipDesign NEEDI (EGAD00001004334), H3Africa H3AChipDesign TrypanoGEN1 (EGAD00001004393), H3Africa H3AChipDesign AWI-Gen (EGAD00001004448), H3Africa H3AChipDesign ACCME (EGAD00001004505), H3Africa H3AChipDesign CAfGEN (EGAD00001004533), H3Africa H3AChipDesign MalSic (EGAD00001004557), H3Africa AWI-Gen Phase 1 WGS data from 100 South Africans (EGAD00001006418).

This study makes use of data generated by the Malaria Genomic Epidemiology Network (MalariaGEN)^13^. A full list of the investigators who contributed to the generation of the data is available from www.MalariaGEN.net. Funding for this project was provided by Wellcome Trust (WT077383/Z/05/Z) and the Bill & Melinda Gates Foundation through the Foundation of the National Institutes of Health (566) as part of the Grand Challenges in Global Health Initiative. The follow MalariaGEN datasets were included in the analysis: whole genome sequencing and genotyping of samples from Mossi from Burkina Faso (EGAD00001005011), Bantu from Cameroon (EGAD00001005012), Semibantu from Cameroon (EGAD00001005013), Chagga from Tanzania (EGAD00001005015), Pare from Tanzania (EGAD00001005016), Wasaamba from Tanzania (EGAD00001005014).

The study also included in-house data generated from 8,463 participants that had not previously been analyzed in this way (South Africa - SABR^14^ [EGAD50000001477], Madagascar^15^ [EGAD50000000708], South Africa - ARK^16^, Aotearoa New Zealand^17^, French Polynesia^18^, Uganda, Indonesia, New Caledonia, and Chile). Participants in these studies were required to be at least 18 years old and willing and able to consent. All participants completed informed consent forms approved by relevant ethics review boards and governing agencies in each country of origin. Where necessary, approvals were obtained for use of controlled-access data (e.g., H3Africa). A full list of cohorts, number of individuals, and source of genomic DNA is provided in Supplemental Table S12. In most cases, genomic DNA originated from whole blood, in some cases from saliva, or, e.g. in the case of 1kGP, from lymphoblastoid cell lines. All sequencing was performed on Illumina platforms with read lengths between 100–150 bp.

### Processing of Sequencing Data and Basic QC

Data was processed with an in-house pipeline using GATK Best Practices. We excluded individuals with excessively low effective coverage (<3×) or evidence of contamination (VerifyBamID2 freemix >0.05). Effective coverage is calculated as [Coverage * (1 - Duplicate Rate) * (1 - Contamination)].

### Allele Frequency Net Database (AFND) frequencies

Allele frequencies and content reported in this paper were aggregated using code from the following github repository: https://github.com/slowkow/allelefrequencies.git using a version of the website which was available on July 6th, 2026. Frequencies reported are derived from the gold standard datasets deposited to the AFND, but a table correlating the gold-standard to all derived allele frequencies is available as Supplemental Table S13.

### Subcontinental group assignment outside of continental supergroup

There were two instances of assignment of subcontinental group outside of the direct continental subgroup affecting 216 individuals, the vast majority affecting samples in the 1kGP or HGDP. 116 African Caribbean in Barbados (ACB) and 74 African Ancestry in USA SW (ASW) assigned to the subcontinental North America group and continental Africa as well as 26 individuals in HGDP from the Mozabite population of Mzab, Algeria assigned to the Northern African group and continental Asia. In each of these assignments, we sought to preserve continental grouping as a method of ancestry evaluation and subcontinental grouping as a reflection of the modern day location of these individuals.

### Inclusion of 1kGP children

Frequencies reported in the manuscript reflect allele frequencies for groups with the 608 children of trios included in the frequencies. While this relatedness skews the sampling design and population representation, we note minimal quantitative changes between the allele frequencies discussed in detail throughout the manuscript both with and without the inclusion of trios. A table with group counts after removal of the children of trios can be found at Supplemental Table S14. A table with allele frequencies both with and without these 608 children can be found in Supplemental Table S30, S31, and S32.

### Benchmarking Design

Algorithms were evaluated on their ability to accurately genotype across all loci in a gene family, performance at various sequencing coverage depths, computational cost, Mendelian consistency, and agreement with orthogonal datasets. Benchmarking genotyping accuracy across such diverse and divergent regions and against sequence-resolved whole genome assemblies necessitated development of a broadly applicable method for comparison, hap-eval (see below).

### HLA Benchmarking (Gold Standard)

HLA typing accuracy on HLA-A, B, and C was evaluated using a recent study of HLA genotypes using targeted exome sequencing^19^. A previous study which leveraged Sanger-derived genotypes^20^ was also referenced, but due to its incomplete and noisy nature, was difficult to integrate. Results for the comparison to the 2014 study are in Table S15. We compared three HLA genotyping tools (HLA*LA^21^, Kourami^22^, and T1K^23^) against the 2018 exome sequencing results at simulated depths of 5×,10×, and 30× for 330 samples from 1kGP. Recall and precision were calculated based on the sample-matched allele calls at each locus. False positives are alleles which are in our set, but not in the gold standard set. False negatives are alleles which are in the benchmarking set, but not in our set. True positives are alleles matched between both sets. Both HLA*LA and T1K performed well with F1-scores above 0.73 at HLA-A, -B, -C and DRB1. At 5× coverage, T1K consistently outperformed HLA*LA across HLA-A, B, and C with an F1-score of 0.80 compared to 0.76 for HLA*LA. Kourami displayed high recall (> 0.73) but due to a large number of ambiguous genotype calls suffered from low precision which drove the overall F1-score down. At 30×, T1K exhibited the highest F1-score across all regions at 0.805 which is largely driven by a Recall of 0.968. Full results for this analysis can be found in Table S16. Raw calls for all genotyping methods tested are found in Supplemental Tables S17,S18,S19. The HLA database used for T1K can be found on Zenodo: Supplemental Material S1.

### Evaluation of accuracy versus haplotype-based assemblies (hap-eval)

Benchmarking for HLA, IGHC, and Fc Gamma Receptor gene families was conducted against haplotype-resolved assemblies produced by the Human Genome Structural Variant Consortium (HGSVC)^24^, Human Pangenome Reference Consortium (HPRC)^25^, and the Platinum Pedigree project^26^. Predicted genotypes in each region were converted back to sequence resolution predictions which were then aligned back to a common reference genome (GRCh38). Comparable genomic regions were then extracted from the alignment of the assemblies to the same reference genome and alignments were compared against each other via CIGAR strings. With this, false positives (variants relative to GRCh38 in the prediction, but not the assembly), false negatives (variants in the assembly, but not in the prediction), true positives (variants in both the assembly and the prediction), and true negatives (non-variant bases aligning to GRCh38 in the assembly and the prediction) could be determined for each aligned base. Full results can be found in Supplemental Table S2.

### Determining the effect of sequencing depth of coverage on number of newly observed alleles

Due to the wide range of sequencing depths of coverage included in the entire cohort, we assessed whether low sequencing coverage generates spurious calls that would incorrectly increase the number of newly observed alleles. To test this, we downsampled the 2,662 individuals from LASI-DAD (part of the South Asian group) to an average depth of coverage of 5× and incorporated those into the results for the rest of the cohort as if these downsampled results were the original calls for this subset.

We selected LASI-DAD for two reasons. First, it is closely size-matched to the Eastern African group (n = 2,593), which contributes the largest number of newly observed alleles (402), so the two are directly comparable without normalization; the downsampled depth of 5× is also comparable to the median depth of the Eastern African group (5.6×). Second, LASI-DAD yields few newly observed alleles at full depth despite its size, so the expected true signal is near zero and alleles appearing only after downsampling can be attributed to depth rather than to genuine group-specific diversity. This makes the comparison maximally sensitive to depth-driven artifacts. We note that this low baseline is consistent with India already being well represented in the AFND, though the argument below depends only on the observed baseline, not on its cause.

Originally, we reported 37 alleles which are group-specific to South Asia and for which no frequency information was available in the AFND across HLA-A, -B, -C, and -DRB1. After downsampling, we report 32 such alleles (Supplemental Figure S2A), of which 22 were shared with the full-depth set. The 15 alleles called only at full depth are largely explained by the downsampled data more frequently reporting a single allele per locus rather than two (Supplemental Figure S2B). Ten newly observed alleles were called only in the downsampled set; these fall within a total of 20 alleles of any kind called only in the downsampled set (Supplemental Figure S2C).

Taking these 10 as an upper bound on depth-driven artifacts in a cohort of ∼2,600 samples, low coverage could account for at most ∼2.5% of the 402 newly observed alleles in the size-matched Eastern African group. For low coverage to generate the Eastern African signal, the rate of depth-driven artifacts would need to be roughly 40-fold higher there than observed here at matched sample size and comparable depth. We therefore interpret the group differences in newly observed alleles as reflecting representation in the AFND rather than sequencing depth: the AFND contains over 50,000 individuals from India alone, compared with just over 1,000 from the entire continent of Africa.

### IGHC Benchmarking (Gold Standard)

IGHC benchmarking was conducted for T1K against published sequences for a selection of 1kGP individuals which were sequenced and assembled using long read sequencing (LRS)^27^ and hap-eval. Results are summarized in Table S20.

### Genotyping of HLA Loci

We genotyped classical HLA class I (A, B, C), class II (DPA1, DPB1, DQA1, DQB1, DRB1, DRB3, DRB4), non-classical HLA class I (E, F, G), and other MHC loci (MICA, MICB, TAP1, TAP2) using two different tools: T1K and HLA*LA. Additional HLA loci were called with T1K only (DMA, DMB, DOA, DOB, DPA2, DPB2, DQA2, DQB2, DRA, DRB5, H, HFE, J, K, L, N, P, R, S, T, U, V, W, Y). These tools were run with default parameters. HLA*LA makes use of a pangenome graph with IMGT nomenclature which can be found here: http://www.well.ox.ac.uk/downloads/PRG_MHC_GRCh38_withIMGT.tar.gz and T1K took as an input IMGT coding sequences for each locus. Each coding sequence was padded with 200 base pairs of intronic sequence and joined with N’s where intronic gaps were not spanned. T1K was used as primary HLA typing method, whereas HLA*LA served as a secondary method for cross-validation.

### Genotyping of IGHC

We genotyped IGH constant regions across the genes IGHG1, IGHG2, IGHG3, IGHG4, and IGHGP using T1K and a custom database (described below) encompassing IMGT alleles and novel alleles detected in HPRC haplotype-resolved genome assemblies.

### Custom IGHC Database

The database for IGHC was created using the IMGT published alleles which can be found here: https://www.imgt.org/IMGTrepertoire/index.php?section=LocusGenes&repertoire=genetable&species=human&group=IGHC in addition to the unique alleles which were present in the 232 HPRC Release 2 assemblies (https://humanpangenome.org/hprc-data-release-2/) and hs1 reference. The IGH genes in the assemblies were identified via liftoff^28^ with the method outlined in the HPRC marker paper^25^ with a sequence identity threshold of 90%. Coding sequences were compared to determine unique alleles and number of unique alleles for each locus are noted in Supplemental Table S21. Each coding sequence was padded with 200 base pairs of intronic sequence and joined with N’s where intronic gaps were not spanned. The custom IGHC database is available on Zenodo: Supplemental Material S2.

### Integration of Novel Allele Calls

T1K claims the ability to produce novel allele calls via single nucleotide variants relative to the alleles in the given database. The primary callset only integrates these novel calls when they result in an exact match to the coding sequence of any other IMGT allele and an absence of a match to the original predicted allele’s coding sequence. While this analysis allowed for calling of alleles which were added in a newer IMGT release (v3.63 vs v3.59), novel calls were also integrated if they resulted in a match to another allele which was already in the current database. The validation of this method can be found below in the section “Validation of Novel Allele Calls”. A summary of incorporated, novel allele calls can be found here: Supplemental Table S22. All novel allele calls and their counts can be found in Supplemental Table S23.

### Validation of Novel Allele Calls

Novel allele calls made by T1K were incorporated if they produced an exact match in the coding sequence of another IPD-IMGT/HLA published HLA allele. Our genotyping graph given to T1K relied on alleles from the v3.59 (2025-01) release and we validated any allele calls which were a match to the coding sequence released in the v3.63(2026-01). As confirmation of this methodology, we looked for presence-absence and geographic distribution of one specific allele: HLA-E*01:152. This allele was first published in May 2025^29^ and was observed in low frequency in Southern, Central, and East Africa. GAIA analysis revealed 27 instances of this specific allele (Supplemental Figure S6). All but two of the observations reported in GAIA occurred in Southern, Central, and Western Africa with the other two occurring in South America.

### Genotyping of Fc Gamma Receptors

We genotyped genes in the Fc Gamma Receptor gene family using Locityper and a pre-release version of the HPRC v2.0 graph. FCGR1A, FCGR1BP, FCGR2A, FCGR2B, FCGR2C, FCGR3A, FCGR3B, and FCGBP were genotyped in GRCh38 coordinates using the following graph: https://s3-us-west-2.amazonaws.com/human-pangenomics/pangenomes/scratch/2025_02_28_minigraph_cactus/hprc-v2.0-mc-grch38/hprc-v2.0-mc-grch38.raw.vcf.gz. Due to an exceedingly large variant spanning the FCGRT locus in the GRCh38-based graph, this gene was genotyped in the hs1-based graph using the following resource: s3://human-pangenomics/pangenomes/scratch/2025_02_28_minigraph_cactus/hprc-v2.0-mc-ch m13/hprc-v2.0-mc-chm13.raw.vcf.gz. Both graphs were then processed according to the best practices explained in: https://locityper.vercel.app/input_output. After graph processing, reference k-mer counting, and locus addition, raw reads were then pre-processed and genotyped with default parameters.

### Genotyping accuracy for all loci against HGSVC assemblies (hap-eval)

The study also included in-housefor their overlap with the HGSVC3 assembly set^24^ and availability of raw data. Comparisons were made both gene-body wide and in coding regions. Gene body wide statistics are captured in Supplemental Table S24. Coding sequence results are contained in Supplemental Table S25.

### Benchmarking universal genotyping tools for IGHC and FCGR

We tested Locityper^30^, T1K^23^, and Ctyper^31^ for their general ability to genotype across both IGHC and FCGR genes. Both Locityper and T1K can receive custom databases or graphs which can be applied genome-wide and Ctyper relies on a pre-computed CNV map incorporating sequence-resolved paralogs. Many, but not all, IGHC and FCGR genes appear in this CNV map. Additionally, vg-giraffe^32^ was tested, but runtimes were not feasible for a cohort of this size. While locityper could recover some IGHC genes (IGHG1, IGHGP) with reasonable accuracy (F1-score > 0.75), this analysis revealed that T1K is more capable in these regions while the opposite was true for FCGR. Results for this comparison can be found in Supplemental Table S26.

### Benchmarking whole genome vs whole exome sequencing for all loci

In addition to comparisons against gold standard sets, we compared tools for consistency against orthogonal data types. We took 314 internal samples from a Ugandan cohort with whole genome sequencing (WGS) and whole exome sequencing (WES) available and initiated genotyping for T1K and Locityper across all genes of interest. We counted true/false positives/negatives similar to the gold standard analysis with the WGS serving as the gold standard against which to compare WES. For WGS, 32 genomic loci showed F1-scores above 0.75 with 31 coming from the HLA gene family. Analysis with hap-eval revealed a higher level of concordance across all regions due to small mismatches in sequence affecting precision in hap-eval less dramatically as compared to a direct matching of genotype which would result in a precision score of 0 for a sample. Results for the comparison using hap-eval can be found in Supplemental Table S27.

### Benchmarking Mendelian consistency

Contained in the 330 1kGP samples were 88 trios. The children in these trios were compared against genotype calls in the parents to determine Mendelian consistency across all regions and simulated sequencing depths. Results from this analysis are contained in Supplemental Table S28. Additionally, hap-eval was used to determine actual sequence match between predicted, inherited alleles and parent genotypes. Results for that analysis are found here in Supplemental Table S29.

### Majority Allelic Diversity (MAD)

We introduce majority allelic diversity (MAD) as a measure of allelic breadth within a group. MAD-X is the number of alleles required for X% of individuals in a group to carry at least one allele from the set. Carrier status is taken from observed diplotypes rather than inferred from allele frequencies, so no Hardy–Weinberg assumption is made. This matters in admixed and structured groups, where genotype frequencies depart from the expectation under allele frequencies alone. The set is built greedily. Beginning with all genotyped individuals in the group, we select the allele carried by the largest number of them and remove every individual carrying it from the pool. Carrier counts are then recalculated among the individuals who remain, and the allele carried by the largest number of those is added next. Selection therefore depends on the residual pool rather than on frequency in the group as a whole: an allele that is common but largely confined to individuals already covered contributes little and may never be selected. Alleles are added until the cumulative number of individuals covered, as a fraction of the group’s original size, reaches or exceeds X%. MAD-X is the size of the resulting set. For example, in a group of 100 individuals, suppose 84 carry allele 001. MAD-50 = 1, since 001 alone exceeds the 50% threshold. Removing those 84 leaves 16 individuals; of these, 12 carry 002, bringing cumulative coverage to 96. MAD-90 = 2.

To extend this to a global measure, we define global MAD-X as the number of unique alleles in the union of all subcontinental MAD-X sets — that is, the total number of alleles needed to simultaneously reach the threshold in every subcontinental group. It should be noted that these numbers can be highly skewed by unequal sampling and population sizes; these figures are meant to be starting points for approaching understanding diversity and vaccine development. In order to directly understand the distribution of HLA alleles in a given population, more direct sampling would be required.

### Measuring per locus diversity with MAD-50

Global MAD-50 values represent the total number of unique alleles across all subcontinental MAD-50 sets in GAIA. Across the eight classical loci surveyed by the AFND, DPA1 has the lowest global MAD-50 (4 alleles). Other loci show even lower allelic diversity, with 15 loci showing a global MAD-50 of 1 (Supplemental Table S7). All of these exist outside of the scope of alleles reported to the AFND and this diversity is reported in GAIA for the first time. All of these regions are contained in the HLA family of genes and there are only 9 loci in HLA which show a MAD-50 above 4: A, B, C, DPB1, DQA1, DQB1, DRB1, MICA, and H. All IGHC and FCGR loci we examined had a global MAD-50 greater than 4, however, MAD-50 should not be confused with fixation of specific alleles. For example, at IGHG1 all subcontinental groups show a MAD-50 between 1 and 3, but there are still alleles which are more common in specific subcontinental groups over others as highlighted in the main text and with a global MAD-50 of 8 (Supplemental Figure S3). Among all loci surveyed, FCGBP has the highest global MAD-50 value of 137 followed by FCGR3A and FCGR2C at 118 and 116 respectively.

HLA-B has the highest mean MAD-50 among the MHC genes with a value of 3.72 between subcontinental groups, followed by DRB1, A, and C with mean MAD-50 values of 2.94, 2.44, and 2.33, respectively. Across all MHC loci, the Southern African, North American, South American, and Eastern African subcontinental groups show the highest mean MAD-50 of 1.41, 1.41, 1.39 and 1.37 respectively. Across all IGHC loci, the top-four subcontinental group MAD-50 values are the Central African (2.2), Southern African (2.2), North American (2.2), and Western African (2.0) subcontinental groups. For FCGR loci, the top-four subcontinental group MAD-50 values are seen in South America (17.7), North America (15.9), South Asia (13.2), and Western Africa (11.7). Another metric for understanding allelic diversity is to simply count the total number of unique alleles which are observed in each subcontinental group. Eastern African subcontinental groups have the highest number of total alleles observed across all HLA loci with a mean of 47.4 alleles per locus, which is supported by previous studies^33,34,35,36^. This Eastern African allele diversity is highly concentrated to A, B, and C with 304, 261, and 347 total alleles respectively. Notably, this subcontinental group shows the highest number of alleles at HLA-DPB1 (199), a locus with a median of 34.0 alleles per group.

### Estimating the effect of group size on MAD

In order to determine the effect that group size has on MAD, we ran a series of simulations based on the underlying data to see if trends remained after equalizing group sizes. We sampled with replacement all subcontinental groups down to 30 individuals per group and calculated allele frequency and MAD statistics for each group (Supplemental Figure S7). We repeated this simulation 1,000 times and empirically derived a 95% confidence interval based on the values observed in the simulations. While the exact numbers derived from the simulated MAD analysis did not match the values observed in the GAIA dataset, the conclusions derived from the simulations were the same: HLA-B was the most variable region based on MAD-50 and MAD-90 as well as AMRS, AMRN, and AFRE being the most variable subcontinental groups based on MAD-50 and MAD-90. One exception is made for the ratio of MAD-90:MAD-50 which does not show the same sort order as the observed ratio in the main text. This is likely due to the fact that MAD-90 values in a simulated small group derived from a diverse group are likely to be depressed due to a bottleneck artifact of the simulation. In a diverse group, much of the total frequency mass is spread across many rare alleles. Downsampling is more likely to miss rare alleles entirely and the few alleles that do survive the bottleneck are the ones that were already relatively common. In a less diverse group, most alleles are already at moderate frequency and survive the downsampling more or less intact and the shape of the frequency distribution is better preserved.

### Bootstrapping to produce confidence intervals for allele frequencies and MAD

To obtain confidence intervals for the allele frequencies we resampled alleles with replacement randomly from the pool of observed alleles within each subcontinental group, drawing alleles equal to the original group size. For each of 1,000 bootstrap replicates, the pool of alleles was established and then allele frequency was calculated from the simulated pool. We then derived 95% confidence intervals for both allele frequencies and MAD values using the percentile method, taking the 2.5th and 97.5th percentiles of the resulting bootstrap distributions. Similarly, for MAD we established a pool of individuals equal to the group size and calculated MAD from that pool over 1,000 bootstrap replicates.

### FCGR haplotype of origin

All haplotypes called for FCGR originate from the HPRC v2 graph referenced above which is primarily composed of 1kGP samples (plus HG002 and linear references). We conducted an analysis of the composition of the origin of each subcontinental group’s haplotype coloring them based on the subcontinental group of the 1kGP sample from which the haplotype was first identified (Supplemental Figure S4) this is referred to as the graph-based haplotype of origin. This analysis revealed expected group trends in FCGR2 and FCGR3 where the majority of graph-based haplotypes of origin in each subcontinental group matched the subcontinental group but these trends were not observed in FCGR1.

## Supplemental Figures

**Figure S1.**
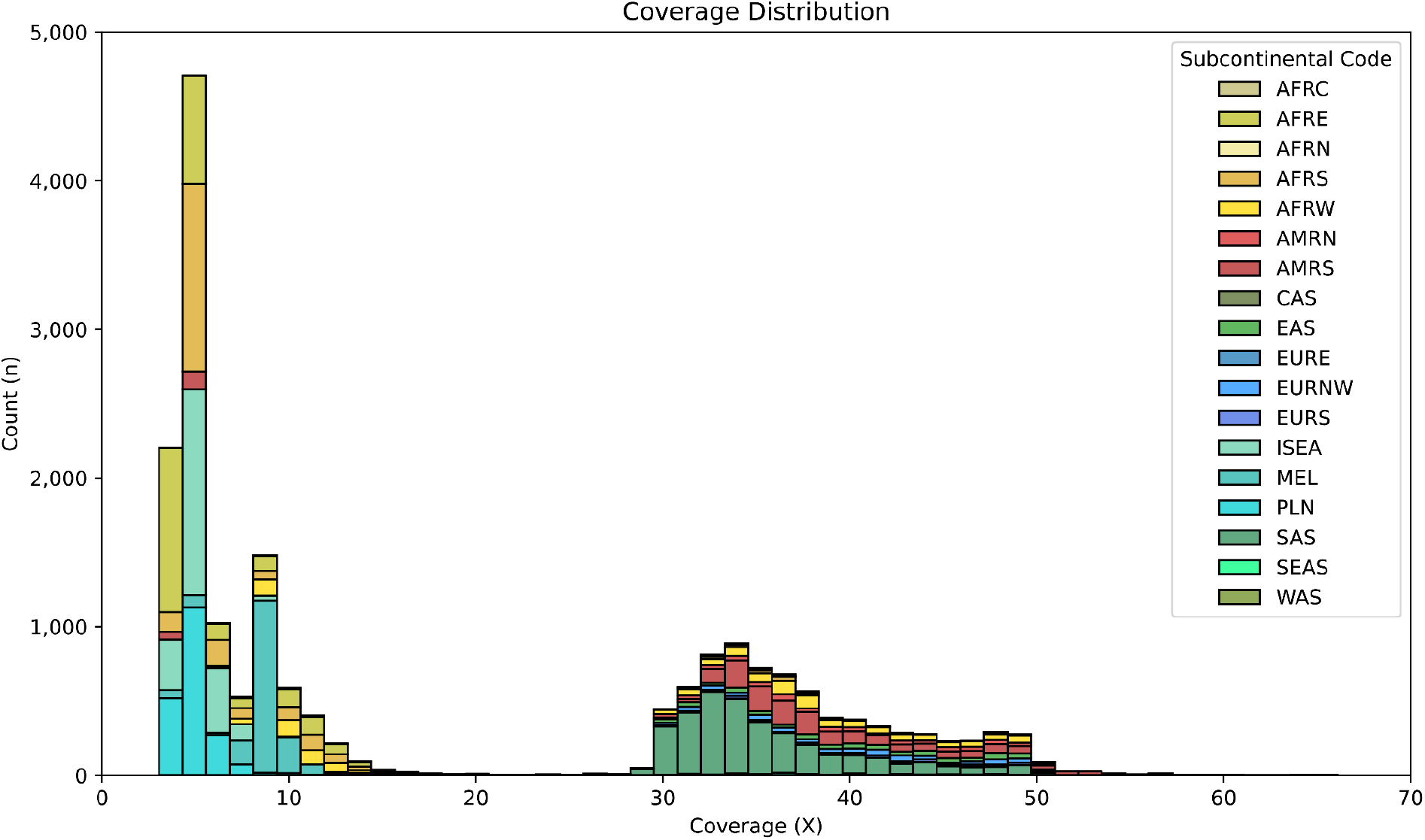
Coverage Histogram for 20,248 individuals in GAIA. Histogram showing sequencing coverage for samples in GAIA colored bar plots indicate subcontinental group.

**Figure S2.**
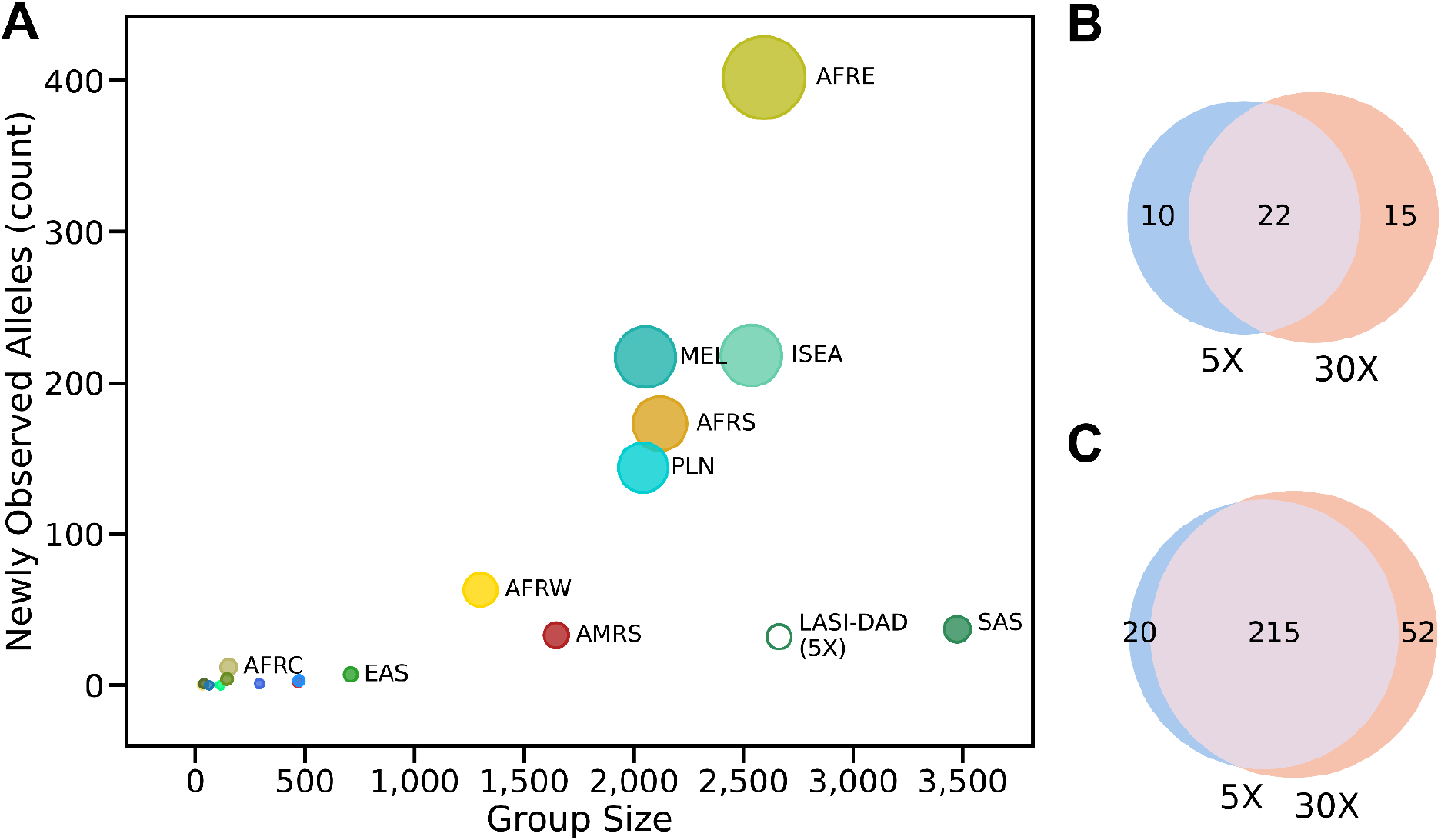
Effect of lower coverage on overall allele discovery. A) Plot showing newly observed allele frequencies compared to the AFND for all original groups and the LASI-DAD downsampled set for HLA-A, -B, -C, -DRB1. B) Venn diagram showing the number of newly observed alleles in the downsampled LASI-DAD set (5X) versus the complete coverage (30X) for HLA-A, -B, -C, -DRB1. C) Venn diagram showing the overall set size and agreement for the total number of unique alleles called with the downsampled LASI-DAD set (5X) versus the complete coverage (30X) for HLA-A, -B, -C, -DRB1.

**Figure S3.**
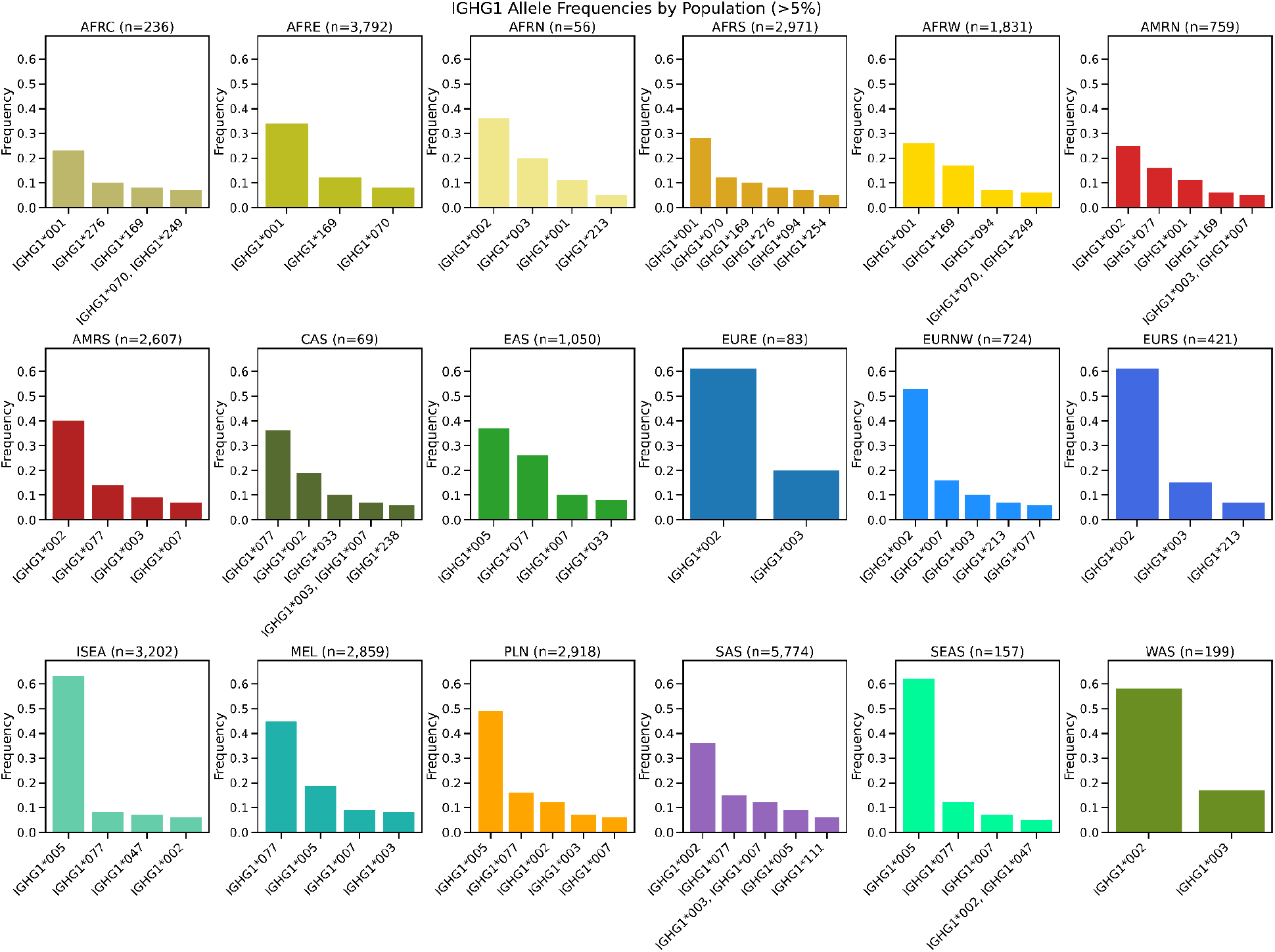
Allele frequency of alleles at IGHG1 which occur at greater than 5% frequency for each subcontinental group. Labels with multiple alleles separated by commas indicate that alleles occur at the same frequency.

**Figure S4.**
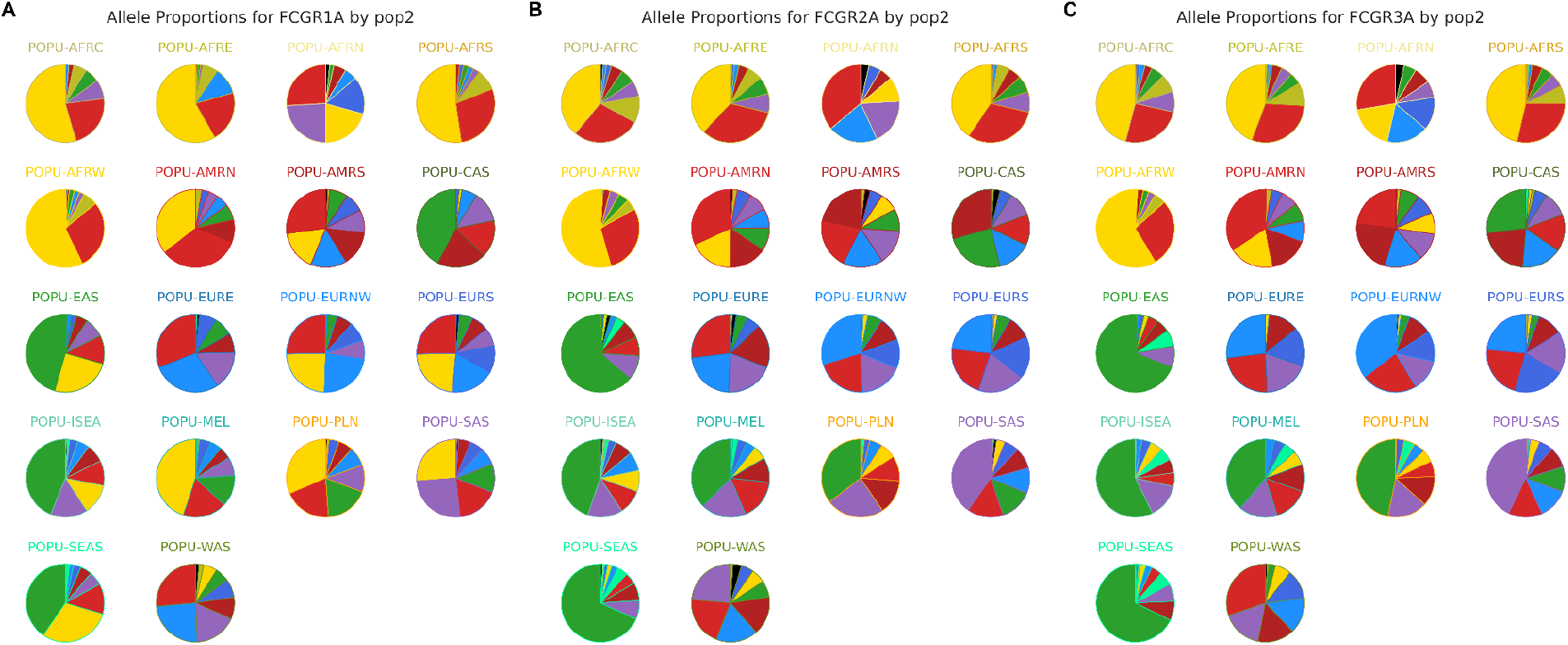
Haplotype of origin assignment for FCGR genes. A) Proportional subcontinental composition of graph originated alleles in each subcontinental group for FCGR1A. Each circle represents all of the haplotypes for each subcontinental group at a given locus. Circles are colored based on the subcontinental designation of the 1kGP genome from which they originated. FCGR1A and other FCGR1 genes show an increase in the number of haplotypes which originate from west African individuals. Specifically, East Asia (EAS), Northwest Europe (EURNW), Southern Europe (EURS), Melanesia (MEL), Polynesia (PLN), South Asia (SAS), and Southeast Asia (SEAS) show this effect to the highest degree. B) Proportional subcontinental composition of graph originated alleles in each subcontinental group for FCGR2A. Each circle represents all of the haplotypes for each subcontinental group at a given locus. Circles are colored based on the subcontinental designation of the 1kGP genome from which they originated. C) Proportional subcontinental composition of graph originated alleles in each subcontinental group for FCGR3A. Each circle represents all of the haplotypes for each subcontinental group at a given locus. Circles are colored based on the subcontinental designation of the 1kGP genome from which they originated.

**Figure S5.**
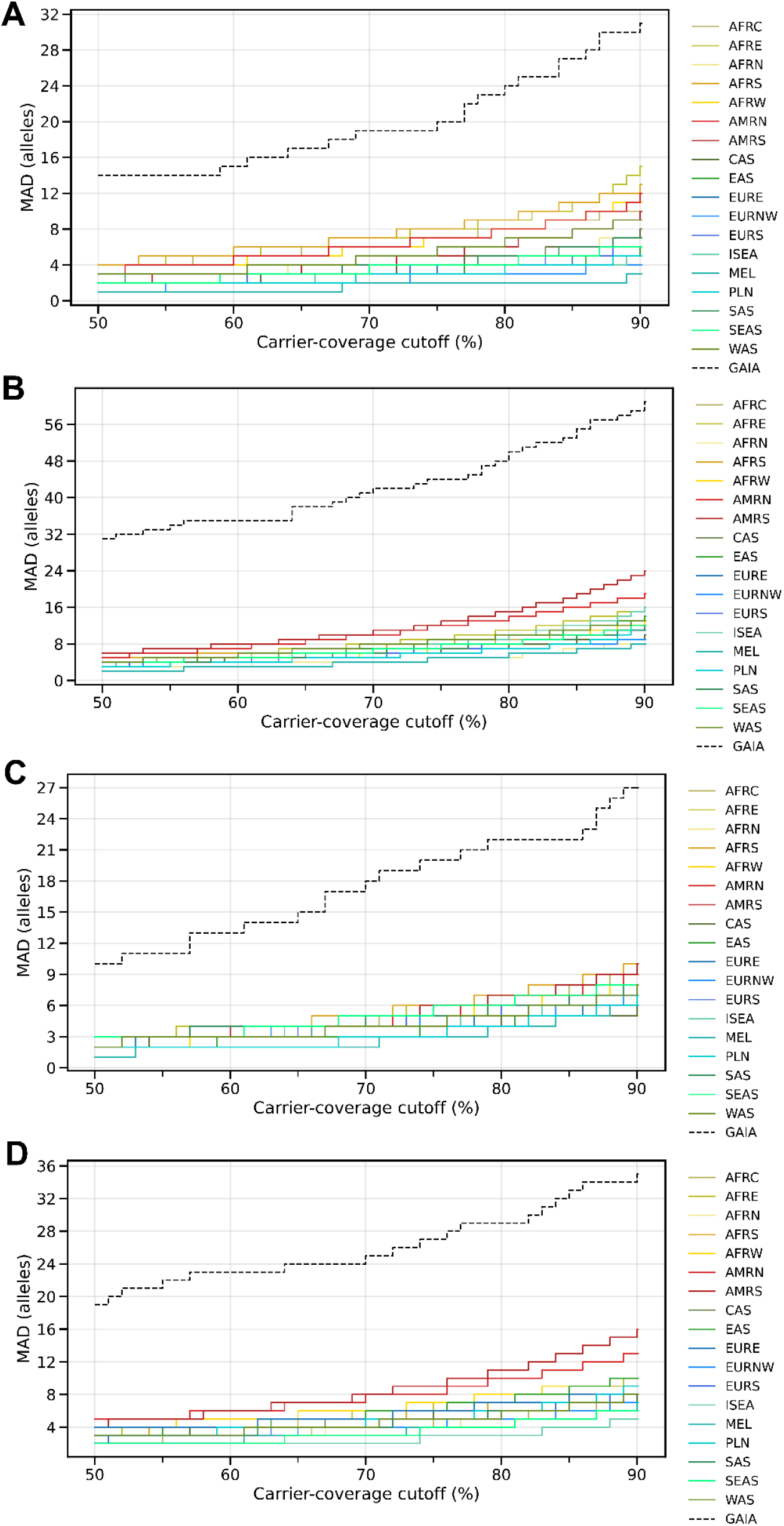
HLA MAD-X. MAD-X plots for HLA-A (A), HLA-B (B), HLA-C (C), and HLA-DRB1 (D) stratified by subcontinental groups and across the entire cohort (GAIA).

**Figure S6.**
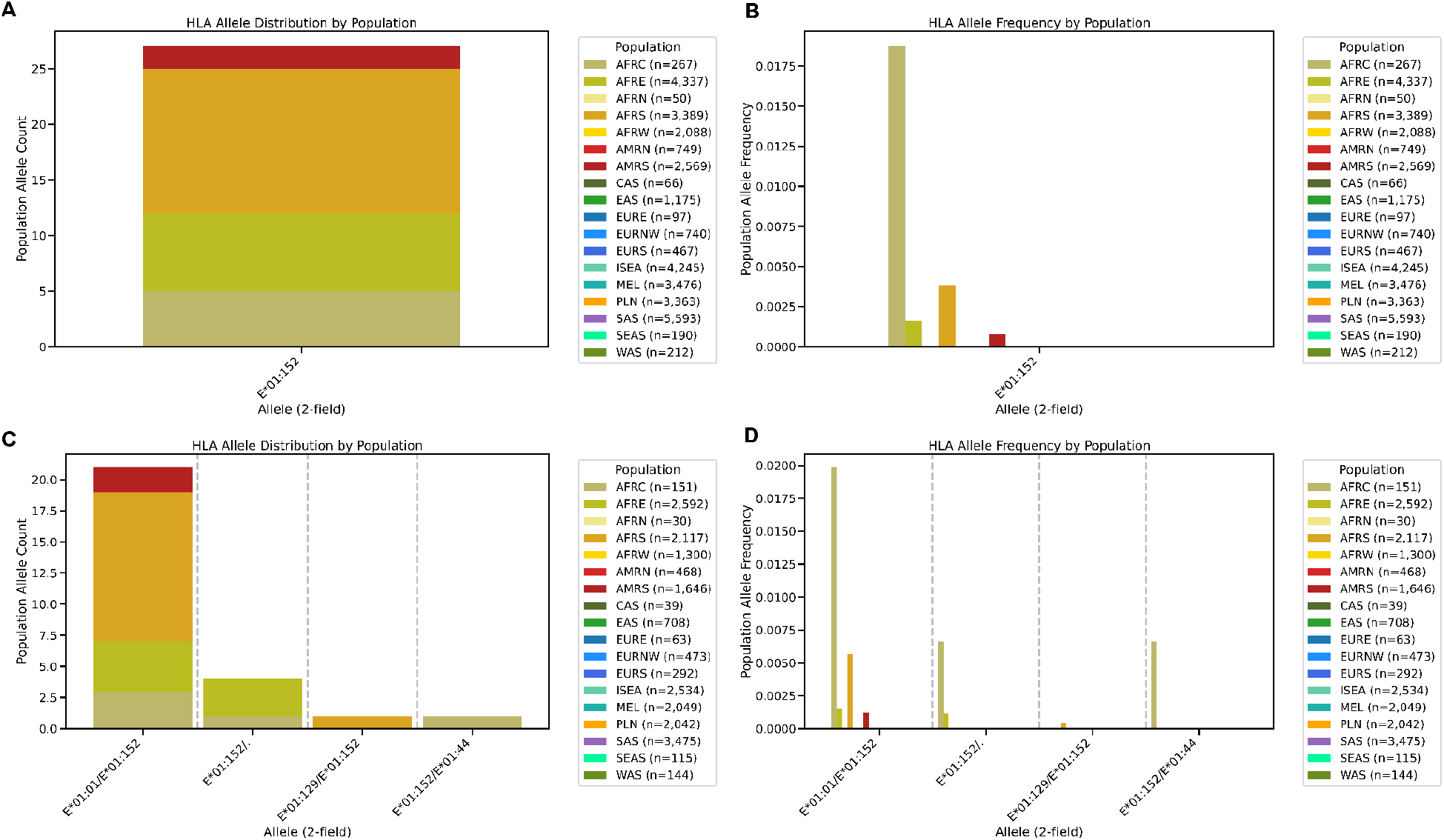
T1K novel-allele calling for HLA-E*01:152. A) Count of HLA-E*01:152 genotypes by subcontinental cohort. B) Allele frequency of HLA-E*01:152 genotypes by subcontinental cohort. C) Count of HLA-E*01:152 alleles by diplotype. Showing which genotypes are inherited together. D) Allele frequencies of HLA-E*01:152 diplotypes.

**Figure S7.**
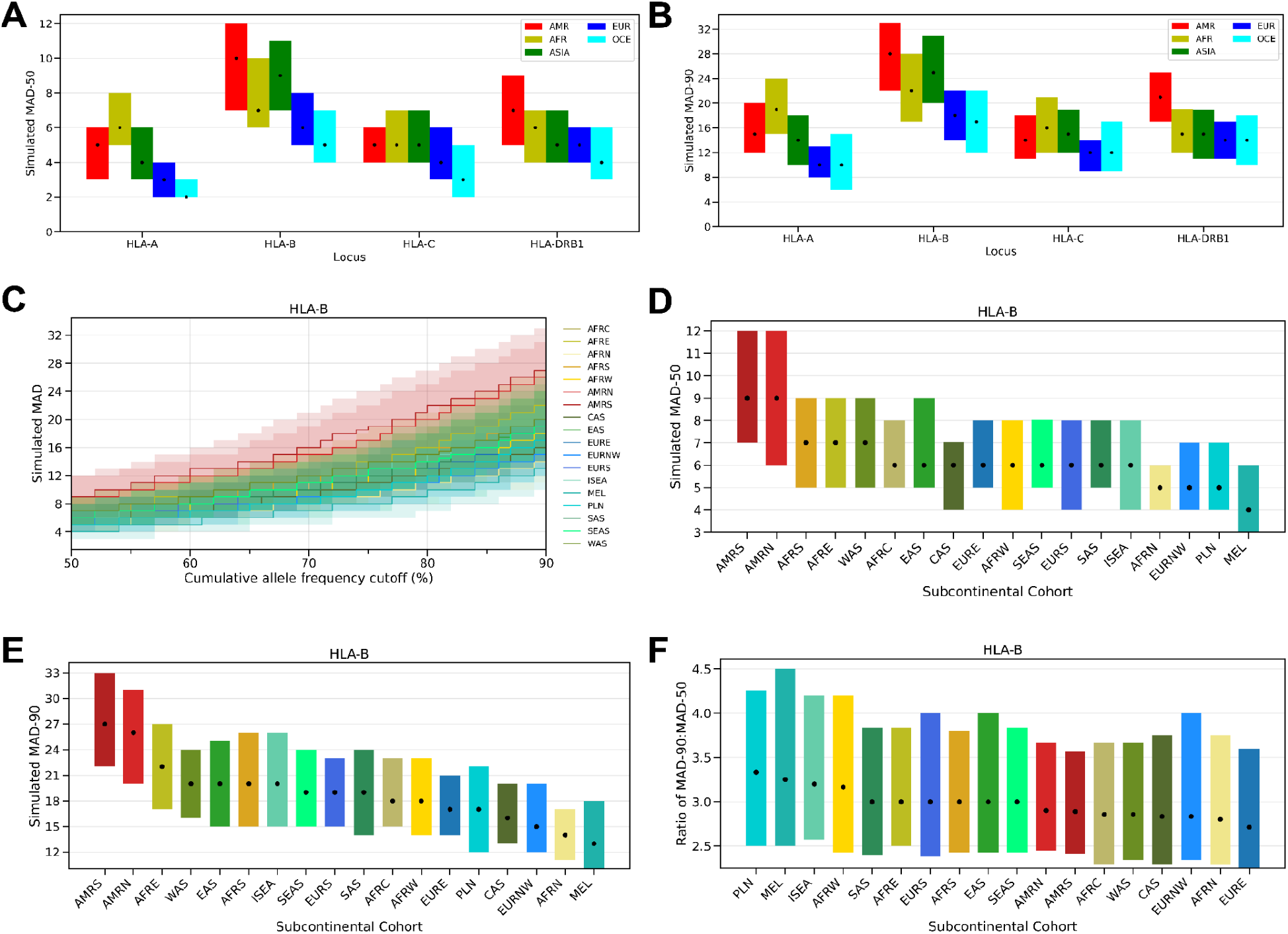
Confidence intervals for MAD calculations derived from sampling with replacement to determine effect of group size. To determine the effect that group size has on MAD, we ran a series of simulations based on the underlying data to see if trends remained after equalizing group sizes. We sampled with replacement all subcontinental groups down to 30 individuals per group and calculated allele frequency and MAD statistics for each group. The middle 95 percentile of outcomes were used to calculate confidence intervals on the number of alleles to guarantee that 50% of individuals in a group have at least one allele in the set (MAD-50, A) or 90% of individuals (MAD-90, B) across HLA-A, -B, -C, and -DRB1 for each continental group. The same analysis was performed at the subcontinental group level for percentages between 50-90 (C), 50 (D) and 90 (E) in addition to F) the ratio of HLA-B MAD-90 to MAD-50 for each subcontinental group.

## Supplemental Tables

**Table S1. Count of individuals in GAIA by subcontinental group. Number of individuals assigned to each of the subcontinental groups in GAIA.** For each group the table lists its short code (e.g., AFRC), the full subcontinental group name (e.g., “Africa, Central”), the individual count, and the modern-day countries whose populations are represented within that group. Columns: Code, Subcontinental Group, Count, Modern Day Countries.

**Table S2. Benchmarking of HLA-, FCGR- and IGHC-region genotyping for 55 1kGP children across sequencing depths (hap-eval).** Precision, recall, and F1 for genotype calls in 55 children of trios, evaluated with hap-eval against haplotype-resolved assemblies from the Human Genome Structural Variation Consortium (HGSVC). Results are broken down per gene (e.g., FCGBP, FCGR1A) and per simulated coverage (5×, 10×, 30×). Columns: Gene, Coverage, Precision, Recall, F1.

**Table S3. HLA alleles whose study-wide GAIA frequency differs from the AFND by more than 5%.** The 24 alleles whose GAIA study-wide frequency differs from the AFND frequency by more than 5 percentage points. For each allele the table reports the GAIA study-wide frequency, the AFND frequency, the raw frequency differential (GAIA − AFND). Columns: Haplotype/Allele, Study-wide Allele Frequency, AFND Allele Frequency, Allele Frequency Differential.

**Table S4. Frequency per subcontinental group of 2,413 alleles reported for the first time by GAIA (all AFND-reported loci).** For each of 2,413 alleles (spanning every locus catalogued by the Allele Frequency Net Database [AFND]) whose frequency information is reported for the first time in GAIA, the observed count within each subcontinental group. Each row is one allele × group observation. Columns: Allele, Group Code, Count.

**Table S5. Frequency per subcontinental group of 1,780 HLA class I/II alleles reported for the first time by GAIA.** For each of 1,780 alleles across HLA-A, -B, -C, and -DRB1 whose frequency information is reported for the first time in GAIA, the observed count in each subcontinental group. Each row is one allele × group observation. Columns: Locus, Group Code (e.g., AFRC), Allele, Count (number of chromosomes on which the allele was observed in that group).

**Table S6. Alleles absent from the AFND but observed at >1% frequency in at least one GAIA group.** The 20 alleles that are not present in the AFND yet reach greater than 1% frequency in a given subcontinental group. For each allele × group pairing the table gives the observed count, the within-group allele frequency, and the study-wide (global) allele frequency, illustrating alleles that are locally common but globally rare. Columns: Haplotype/Allele, Group Code, Count, Group Allele Frequency, Study-wide Allele Frequency.

**Table S7. Majority allelic diversity (MAD) at multiple thresholds for all GAIA groups.** MAD counts computed at thresholds for every 5% from 50 to 95 for every subcontinental group and locus. For each group × locus × cutoff the table gives the number of alleles and allele names needed to reach the threshold. The values are calculated for both with and without 1kGP trios. Columns: Group code, Locus, 1kgp Trio Probands, MAD-X, Allele Count, Alleles.

**Table S8. Clinically actionable HLA alleles with supporting literature.** A literature-review compilation of clinically actionable HLA alleles, included regardless of their frequency in or absence from GAIA. For each allele the table lists the locus, the associated condition(s), population and functional remarks (mechanism, effect direction, population context), and short-form reference(s). Columns: Locus, Allele, Associated Condition(s), Population & Functional Remarks, Reference. Full citations are in Table S9.

**Table S9. Full citations for Table S8. Complete bibliographic references (authors, year, title, journal, volume, pages) for the short-form citations used in Table S8.** One reference per row.

**Table S10. Annotated alleles observed at >1% in one group but <1% in every other group. Alleles present at greater than 1% allele frequency in a single subcontinental (or continental) group and below 1% in all others, annotated with disease/immune associations.** Each row records the group codes, locus and allele, an evidence tier, the association category (e.g., HIV, Autoimmune), the direction of effect (Susceptibility/Protective), and free-text notes summarizing the supporting literature. Columns: Group Code, Locus, Allele, Tier, Category, Effect, Notes.

**Table S11. Counts of continental-to-subcontinental group relationships. Mapping between continental groups and their constituent subcontinental groups with individual counts.** Each row pairs a continental group (e.g., POPU-AFR) with one of its subcontinental groups (e.g., POPU-AFRC) and the count of individuals in that subcontinental group. Columns: Continental Group, Subcontinental Group, Count.

**Table S12. Source and individual counts for all substudies included in GAIA. Per-cohort breakdown of the studies aggregated into GAIA.** For each cohort the table gives its name, whether it is an external or internal source, the number of individuals contributed, and the source genomic DNA material (e.g., LCL, Blood). Columns: Cohort, Source, Number of individuals, Source Genomic DNA.

**Table S13. AFND gold-standard vs all-AFND allele frequencies.** Comparison of AFND allele frequencies computed from all AFND studies versus the AFND gold-standard subset only. For each allele the table gives its locus, the frequency across all AFND studies, the frequency in the gold-standard subset, and the difference (gold − all). Columns: Allele, Locus, AFND AF (ALL), AFND AF (GOLD), DIFF (GOLD - ALL).

**Table S14. Subcontinental group counts after removal of 1kGP trio children.** Number of individuals in each subcontinental group after the 1kGP children of trios are removed (i.e., the de-duplicated cohort used for frequency estimation). Columns: Subcontinental Group, Count.

**Table S15. HLA gold-standard benchmarking against Gourraud (2014) for three HLA callers.** Precision, recall, and F1 for Kourami, HLA*LA, and T1K compared against the Sanger sequence-derived HLA genotypes of 1,267 1kGP individuals (Gourraud et al., 2014), reported per locus. “Library Type” indicates the downsampling scenario used to model our study data; LBTP-WGS-30× corresponds to full study-equivalent 30× WGS. Columns: Locus, Precision, Recall, F1, Caller, Library Type.

**Table S16. HLA gold-standard benchmarking against Abi-Rached (2018) for three HLA callers.** Precision, recall, and F1 for Kourami, HLA*LA, and T1K compared against the exome-derived HLA genotypes of 2,693 1kGP individuals (Abi-Rached et al., 2018), reported per locus and caller. “Library Type” indicates the downsampling scenario modeling our study data; LBTP-WGS-30× corresponds to 30× WGS. Columns: Locus, Precision, Recall, F1, Caller, Library Type.

**Table S17. Raw allele calls for the benchmarking set from Kourami for HLA genes.** Raw allele calls from Kourami for 330 1kGP samples inside of which there are 88 children of 1kGP trios at 2 simulated sequencing depths used for HLA gold-standard benchmarking. 30× was omitted due to poor performance at low depths.

**Table S18. Raw allele calls for the benchmarking set from T1K for HLA genes.** Raw allele calls from T1K for 330 1kGP samples inside of which there are 88 children of 1kGP trios at 3 simulated sequencing depths used for HLA gold-standard benchmarking.

**Table S19. Raw allele calls for the benchmarking set from HLA*LA for HLA genes.** Raw allele calls from HLA*LA for 330 1kGP samples inside of which there are 88 children of 1kGP trios at 3 simulated sequencing depths used for HLA gold-standard benchmarking.

**Table S20. IGHC gold-standard benchmarking (T1K) against 105 long-read–resolved 1kGP individuals (Jana, 2025).** Genotyping performance of T1K on the IGHC (IGHG) genes, evaluated with hap-eval against 105 1kGP individuals whose IGHC sequences were resolved by targeted long-read sequencing (Jana et al., 2025). For each gene × coverage the table reports mean true positives, true negatives, false positives, false negatives, observations, and errors, followed by precision, recall, and F1. Columns: Gene, Coverage, True Positives (Mean), True Negatives (Mean), False Positives (Mean), False Negatives (Mean), Observations (Mean), Errors (Mean), Precision, Recall, F1.

**Table S21. Number of unique alleles in the T1K database for IGHC genotyping. Per-gene composition of the T1K IGHC allele database.** For each IGHC gene (IGHG1, IGHG2, IGHG4, IGHGP) the table gives the number of unique alleles from IMGT, the number of novel unique alleles derived from the HPRC Release 2 assemblies, the number of alleles overlapping between the two sources, and the resulting total number of alleles in the database. Alleles from HPRC R2 were compared against existing IMGT alleles per locus and only unique sequences retained. Columns: Gene, IMGT unique alleles, HPRC R2 unique alleles, Overlap, Total alleles in DB.

**Table S22. Genotype changes resulting from T1K novel-allele calling.** Full list of calls in which T1K’s original allele assignment changed to a different, exactly matching allele after applying the single-nucleotide changes T1K identified. For each change the table lists the group codes, the original allele called, the new allele after the SNV(s) were applied, the group level (pop1/pop2), and the count of individuals affected. Columns: Group Code, original_allele, new_allele, level, count.

**Table S23. All novel allele calls and their counts.** All novel allele calls generated by T1K for HLA genes, with observation counts.

**Table S24. Whole-gene-body precision, recall, and F1 for genotype calls (hap-eval).** Per-sample, per-region, per-haplotype benchmarking for 55 1kGP samples at multiple sequencing depths, evaluated with hap-eval against the entire gene body of the HGSVC haplotype-resolved assemblies. Each row reports raw confusion-matrix counts and derived metrics for one sample × region × haplotype. Columns: SAMPLE, SEQ_COV, TP, TN, FP, FN, OBS, ERR, REGION, HAP, PREC, RECALL, F1.

**Table S25. Coding-region precision, recall, and F1 for genotype calls (hap-eval).** As in Table S24, but restricting the comparison to coding regions only of the HGSVC haplotype-resolved assemblies, for 55 1kGP samples across sequencing depths. Each row is one sample × region × haplotype. Columns: SAMPLE, SEQ_COV, TP, TN, FP, FN, OBS, ERR, REGION, HAP, PREC, RECALL, F1.

**Table S26. Cross-tool comparison of FCGR and IGHC genotyping (Locityper, Ctyper, T1K).** Benchmarking of three tools — Locityper, Ctyper, and T1K — across simulated depths, compared against HGSVC sequence-resolved assemblies for 55 1kGP individuals. Precision, recall, and F1 are reported per locus (rows), with grouped column blocks per tool and per coverage: Locityper (5×/10×/30×), Ctyper (5×/10×/30×), and T1K (10×/30×).

**Table S27. WES-vs-WGS concordance for 314 Ugandan individuals (hap-eval).** Precision, recall, and F1 for whole-exome-sequencing (WES)–derived alleles compared against whole-genome-sequencing (WGS)–derived alleles called by the same tool for 314 Ugandan individuals, evaluated with hap-eval. Each row is one sample × region × haplotype. Columns: SAMPLE_INDEX, SEQ_COV, TP, TN, FP, FN, OBS, ERR, REGION, HAP, PREC, RECALL, F1.

**Table S28. Mendelian-inheritance benchmarking matrix for 88 children of trios.** Per-locus Mendelian consistency across all immune gene loci for 88 children of trios. Each cell scores a child’s two predicted alleles against the parental alleles: 2 = both predicted alleles seen in a parent, 1 = exactly one predicted allele matched, 0 = neither matched any parental allele; an allele matching both haplotypes of a single parent is counted once. Rows are individuals; columns are the loci (HLA-A, -B, -C … MICA, MICB, TAP1/2, IGHG1–4, FCGR genes, etc.) plus a coverage column. Columns: locus (sample ID), one column per gene, coverage.

**Table S29. Mendelian-inheritance haplotype precision, recall, and F1 (hap-eval).** Per-sample, per-region, per-parental-haplotype benchmarking for 87 1kGP children of trios across sequencing depths, comparing each child’s genotyping-tool calls against the alleles predicted from the parents. HAP indicates the parental haplotype of origin (e.g., mat/pat). Columns: SAMPLE, SEQ_COV, TP, TN, FP, FN, OBS, ERR, REGION, HAP, PREC, RECALL, F1.

**Table S30.** GAIA allele frequencies with and without 1kGP children included (T1K - HLA). GAIA allele frequencies computed with and without the 1kGP trio children, for T1K HLA calls.

**Table S31.** GAIA allele frequencies with and without 1kGP children included (T1K - IGHG). GAIA allele frequencies computed with and without the 1kGP trio children, for T1K IGHG calls.

**Table S32.** GAIA allele frequencies with and without 1kGP children included (FCGR - Locityper). GAIA allele frequencies computed with and without the 1kGP trio children, for Locityper FCGR calls.

**Material S1. ZENODO.** ighc_t1k_db.tar.gz T1K database for IGHC genes.

**Material S2. ZENODO.** hla_t1k_db.tar.gz T1K database for HLA genes.

